# A multitude of dyes but unsatisfactory staining: Physico-chemical profile and factors associated with staining quality of Romanowsky-type stains used for malaria microscopy in Plateau State, Nigeria

**DOI:** 10.1101/2025.04.07.25325361

**Authors:** Stephen Eghelakpo Akar, David Magbagbeola Dairo, Remi Kehinde, Tosan Erhabor, Mary Ogheneruemu Akar, Godwin Ntadom, Treasure Obianuju Okoye Wariboko, Patrick Nguku, Shakir Balogun, Oladosu Oladipo, Wellington Oyibo, Olufemi Ajumobi, Ikeoluwapo Ajayi

## Abstract

**Introduction:** There is paucity of studies done to assess the composition and staining quality of Romanosky-type stains used for malaria microscopy in Nigeria.

**Methods:** We conducted a cross-sectional study among 92 laboratory facilities and subjected them to UV-spectroscopy, HPLC, GC, ashing and pH measurement. We conducted multivariable binary logistic regression analysis at p<0.05.

**Result:** Of 94 stains, 38(40.4%) were Field, 36(38.3%) were Giemsa, and 20(21.3%) were Leishman stains. Overall, 23.7%, 33.3% and 25.0% of Field, Giemsa and Leishman stains respectively yielded satisfactory staining. Predictors of staining quality were proportion of Azure B>20% (aOR = 15.1, 95% CI = 2.62-115.00); Azure B to Eosin Y ratio >1.5 (aOR = 4.55, 95% CI = 1.03-25.10); and alcoholic solvent (aOR = 27.70, 95% CI = 2.43-462.00).

**Conclusion:** Romanowsky-type stains used for malaria microscopy in Plateau State were of variable composition and mostly lacked the RGE-relevant dyes required for good nuclear and cytoplasmic differentiation of malaria parasites.

## Introduction

Malaria, a mosquito-borne parasitic disease caused by *Plasmodium* species, has remained a major global public health concern (Poespoprodjo et al., 2023). Worldwide, the number of malaria cases was estimated to be 263 million with an incidence of 60.4 cases per 1000 population, accounting for an estimated 597 000 deaths, with a mortality rate of 13.7 per 100000 in 2022. In 2023 alone, the 11 high burden to high impact (HBHI) countries-Burkina Faso, Cameroon, the Democratic Republic of the Congo, Ghana, India, Mali, Mozambique, Niger, Nigeria, Uganda and the United Republic of Tanzania - accounted for 66% of all malaria cases and 68% of deaths globally. Between 2017 and 2023, the incidence of malaria cases in these 11 countries increased by 13.8%, from 152 million to 173 million, and mortality increased by 2.3%, from 399 000 to 408 000. (WHO, 2024). In 2022, the African Region bore the heaviest malaria burden, with 94% of cases and 95% of deaths globally, representing 233 million malaria cases and 580 000 of deaths, a small reduction compared to 2021(WHO Regional Office for Africa, 2025). Nigeria accounts for the highest incidence of global malaria burden compared to any other country, with 27% of the global estimated malaria cases and 31% of the estimated deaths, as well as an estimated 55% of malaria cases in West Africa in 2022. Nigeria’s large population, combined with other factors such as sanitation and ecological characterisitcs that favor mosquito breeding, account for the endemicity of malaria transmission (Severe Malaria Observatory, 2025). WHO has recommended prompt malaria diagnosis either by microscopy or rapid diagnostic tests (RDTs) for all patients with suspected malaria before administering treatment. Early and accurate diagnosis is essential both for effective case management and for malaria surveillance. Parasite-based diagnostic testing significantly reduces illness and death by enabling health providers to swiftly distinguish between malarial and non-malarial fevers and select the most appropriate treatment. It improves the overall management of patients with febrile illnesses and may also help reduce the emergence and spread of drug resistance (WHO, 2025). Although the introduction of rapid diagnostic tests (RDTs) has demonstrated the potential to ameliorate some diagnostic limitations associated with light microscopy, RDT kits are still faced with challenges such as inability to quantify parasite load for patient treatment monitoring and follow up (NMIS, 2015). Hence, Giemsa stain microscopy has remained the gold standard for routine parasite-based malaria diagnosis in resource-poor countries (Idowu et al., 2015; Smith et al., 2001). However, Mokuolu et al. (2016) have questioned the higher positivity rate of malaria diagnosis by microscopy in Nigeria compared to RDT as they opined that higher prevalence due to microscopy does not reflect the progress and gains that have been made by extant control efforts (Mokuolu et al., 2016). Microscopy involves the staining of either thick or thin blood films with the Romanowsky-type stains such as Giemsa, Leishman or Field stain, however, the WHO recommends Giemsa stain (WHO, 2016a). It has been shown that there is a strong correlation between the physico-chemical properties of these stains and their performance when applied to blood smears (Friedrich et al., 1990; Kraft, 1993; Marshall, 1978a; Wittekind, 2002). The staining process follows an ordered chemical interaction between stain components and cellular constituents in blood (Horobin, 2011; Horobin and Walter, 1987a; Wittekind and Gehring, 1985). Unlike malaria RDT kits, there is no known program for evaluating the quality of microscopy reagents like the WHO-FIND program that certifies RDT kits (Ashraf et al., 2012; Harvey et al., 2017; WHO-FIND, 2014). In North America, the Biological Stains Commission certifies biological stains for diagnostic use, but such is non-existent in WHO Africa Region. Evidence from the field shows that many facilities source their microscopy reagents, including stains from the open market. The supply of in vitro diagnostic reagents and equipment for microscopy and other laboratory tests is poorly regulated in developing countries, where evidence of stocking, sales and supply of sub-standard diagnostic products has been demonstrated (Mori et al., 2011; Engel et al., 2016; Rugera et al., 2014; McNerney and Peeling, 2015). The accuracy of malaria microscopy depends on maintaining a high level of staff competence and performance, ensuring good-quality reagents and equipment at all levels and regular external quality assessment (Garba et al., 2016; Munene et al., 2017; Ghai et al., 2016; WHO, 2016b). Microscopy stain solutions are either prepared by laboratory personnel locally using component dye powders (in-house stains) or purchased as ready-to-use aqueous stock solution (commercially prepared). The quality of the Romanowski-type stains used for malaria microscopy cannot be guaranteed if their physico-chemical properties are compromised (Horobin, 2011; Marshall et al., 1978, 1975a; Penney et al., 2002; Wittekind, 2002). Poor quality stains have the potential to compromise the quality of malaria microscopy, even if personnel are competent and have access to efficient equipment (Marshall et al., 1978, 1975a). Poor quality microscopy could result in either over diagnosis or under-diagnosis of cases with an attendant unnecessarily increase disease prevalence, over prescription of ACTs, and increase in drug resistance. Under-diagnosis of cases, on the other hand, may lead to under-reporting of cases and increased morbidity and mortality (Altaras et al., 2016; Kahama-maro et al., 2011; Miller and Sikes, 2015). In most secondary and tertiary facilities, microscopy continues to be the preferred method for parasite-based diagnosis of malaria (Mokuolu et al., 2016). However, internal quality assurance procedures tend to vary between facilities with an attendant variation in test results when compared from laboratory to laboratory. Reagents for diagnostics are sourced from various sources in the open market, a large proportion of which do not fulfill regulatory requirements for the importation, stocking, storage and supply of in vitro diagnostics (MLSCN, 2018). The quality of staining quality by the Romanowski-Giemsa stains depends on a number of factors, including stain composition, both in the solid and liquid states, which are related to the physico-chemical properties of the stain, manner of constitution for use, storage conditions and duration of time of applying it to the biological material (Penney et al., 2002; WHO, 2016c). The Romanowski-Giemsa staining process follows an ordered chemical reaction in which the dye components interact with each other and cellular components of biological materials. There is therefore, a correlation between the physico-chemical properties of the stains and their staining quality (Friedrich et al., 1990a; Friedrich and Seiffert, 1990; Horobin, 2011; Horobin and Walter, 1987b; Marshall et al., 1975b; Marshall, 1975a, 1975b; Wittekind and Gehring, 1985a). A major problem encountered in the routine use of commercial Romanowsky-type stains is batch-to-batch variations in staining properties, which is usually caused by variations in dye composition and the presence of contaminating metallic salts (Marshall et al., 1975b; Marshall, 1974). Stains standardization is therefore necessary to ensure consistently good stain performance and to facilitate stained slides comparison between laboratories (Horobin, 2011; Marshall et al., 1975a; Schulte, 1991; Scott and French, 1924). Uniformly stained blood smears reduce diagnostic errors and allow morphological features to be correctly identified either by reference to atlases or slide comparison (Khan et al., 2011; Maguire et al., 2006; Marshall et al., 1978, 1975c). Liquid chromatographic technique such as thin layer chromatography (TLC) and high-performance liquid chromatography (HPLC), uv-spectrophotometry, sulphated ash percent determination, and staining test have been demonstrated to be the most effective techniques for quality control of stains (Horobin and Murgatroyd, 1967; Marshall et al., 1975b; Marshall, 1976a; Marshall and Lewis, 1974; Proctor and Horobin, 1985; Galbraith, 1980). Unfortunately, these techniques appear not to be employed by most commercial manufacturers of biological stains to ascertain the quality of their products (Gordon B. Proctor and Richard W. Horobin, 1985; Horobin and Walter, 1987b; Krafts et al., 2010; Wittekind, 2002). By using these techniques, the specifications of successful Romanowski-type stains such as Giemsa and Leishman have been determined. The simplest commercial stains were found to comprise Methylene Blue, Azure B and Eosin Y Horobin, 2011; Marshall, 1976; Penney et al., 2002). The presence of certain other dyes, such as Azure A, sym-Dimethylthionine, Azure C, thionine, 2’,4’,5’-tribromofluorescein, (Fluorescein), although not necessary for the Romanowsky-Giemsa effect, have been shown not to have a detrimental effect on stain performance and characteristics; whereas the presence of methylene violet Bernthsen, methyl thionoline and high concentrations of metallic salts have been proved to be harmful to stain performance as regards the Romanowsky-Giemsa effect (Marshall, 1978b; Marshall et al., 1978; Roe et al., 1941). The objective of the study, therefore, was to describe the physico-chemical characteristics of Romanowski-type stains used for malaria microscopy in Plateau State and determine factors that affect the quality of colour imparted on malaria parasites in stained blood smears. To our knowledge, this is the first attempt to conduct this kind of study in Nigeria.

## Results

### General Stain Characteristics

Of the 94 stains samples, 40.4% (38/94) were Field stains, 38.3% (36/94) were Giemsa stains, 21.3% (20/94) were Leishman stains, and 1.1% (1/94) was a reference stain (Marshall and Lewis stain). Overall, 21.3% (20/94) of stains were manufactured by BEMA Scientific (Field stain: 28.9%; Giemsa stain: 16.7%; and Leishman stain: 15.0%). Batch number was indicated on the bottles of 43.6% (41/94) of all stains (Field stain: 21.1%; Giemsa stain: 63.9%; and Leishman stain: 50.0%). Expiry date was indicated on 42.6% (40/94) of all stains (Field stain: 21.1%; Giemsa stain: 63.9%; and Leishman stain: 45.0%. Commercially prepared stains accounted for 68.1% (64/94) of all stain solution (Field stain: 86.8%; Giemsa stain: 47.2%; and Leishman stain: 70%). The location (company address) of stain manufacturer was known for 68.1% (64/94) of all stains (Field stain: 63.2%; Giemsa stain: 72.2% and Leishman stains: 70.0%) (**Table 1**).

**Table 1.**
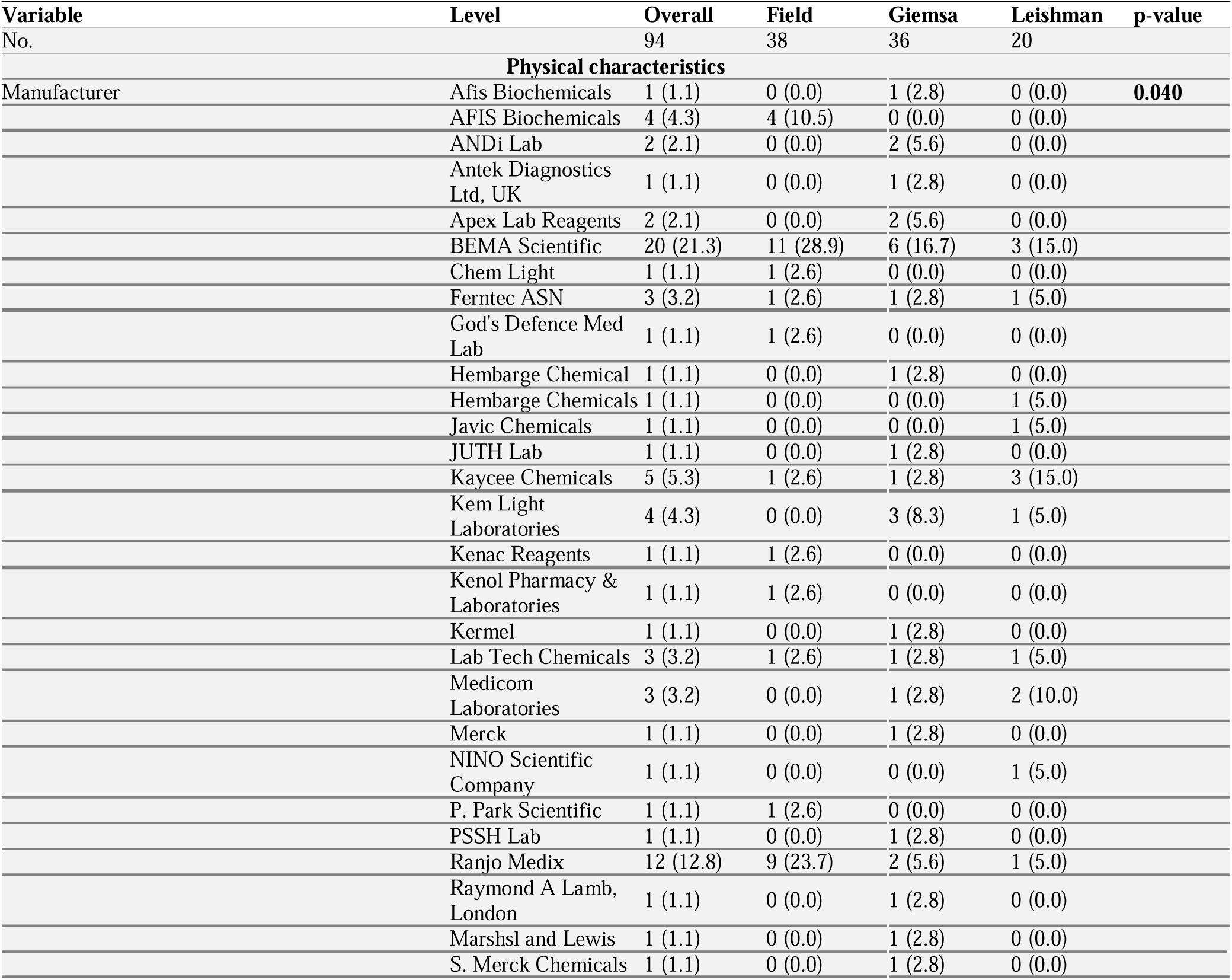
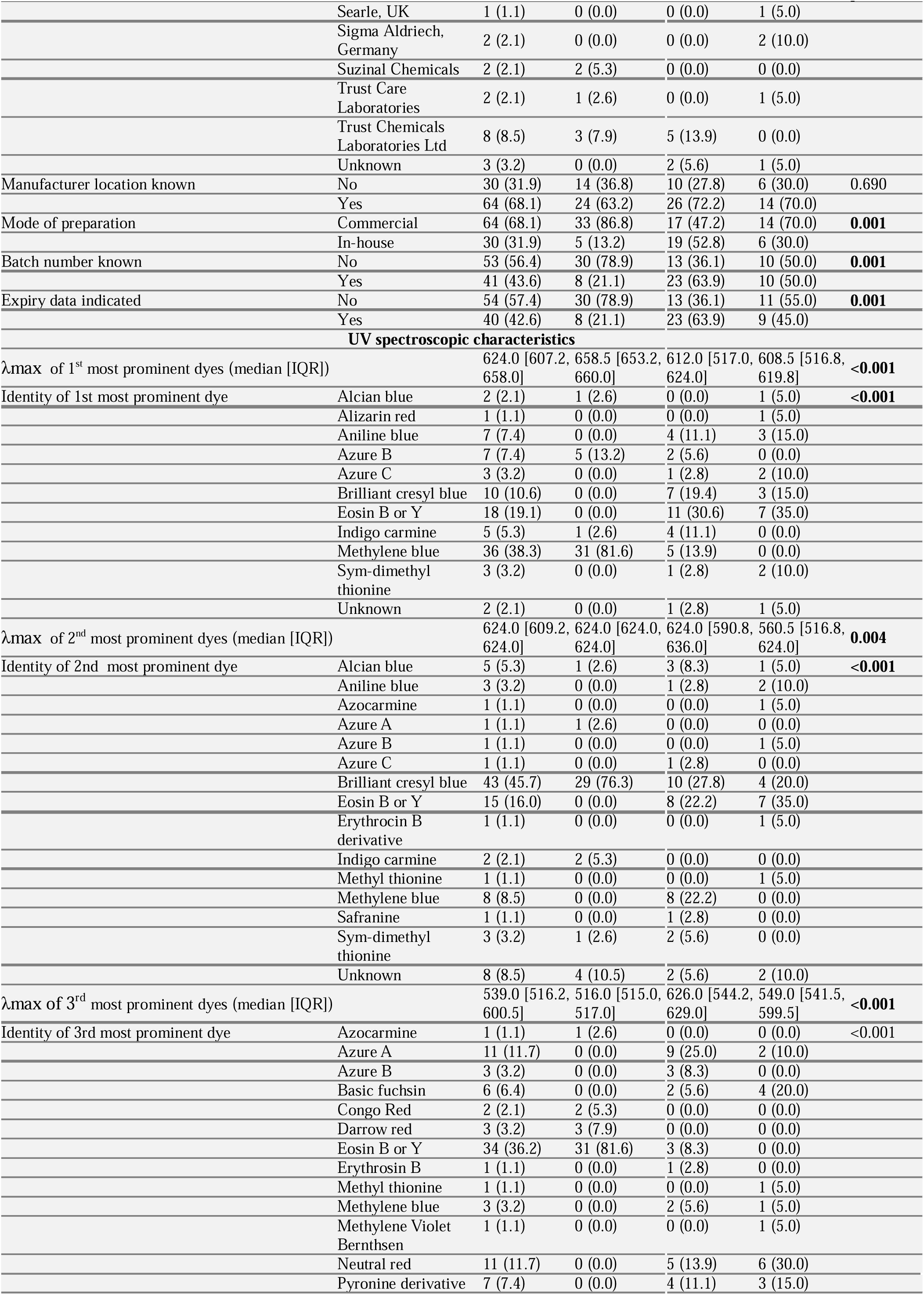
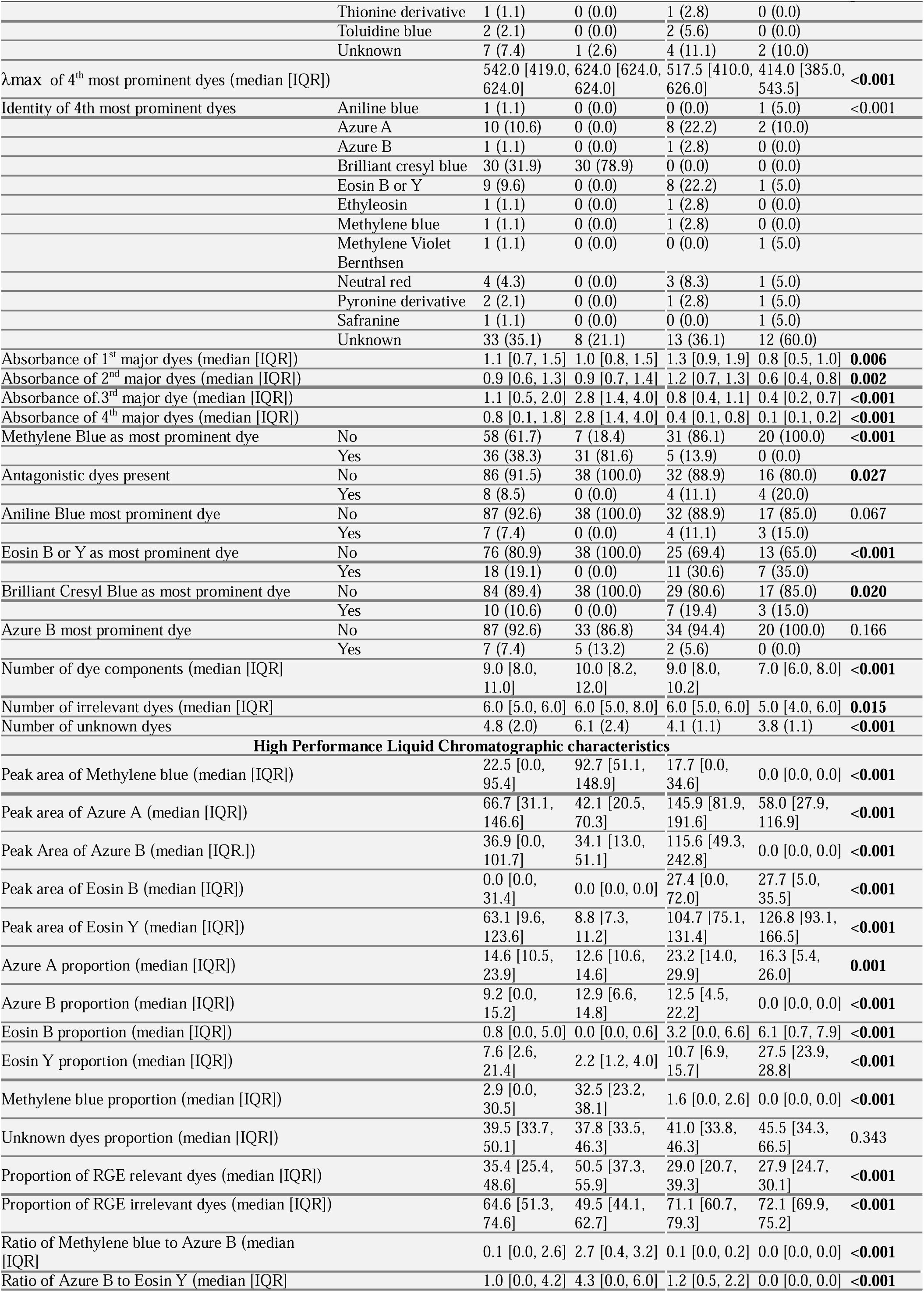
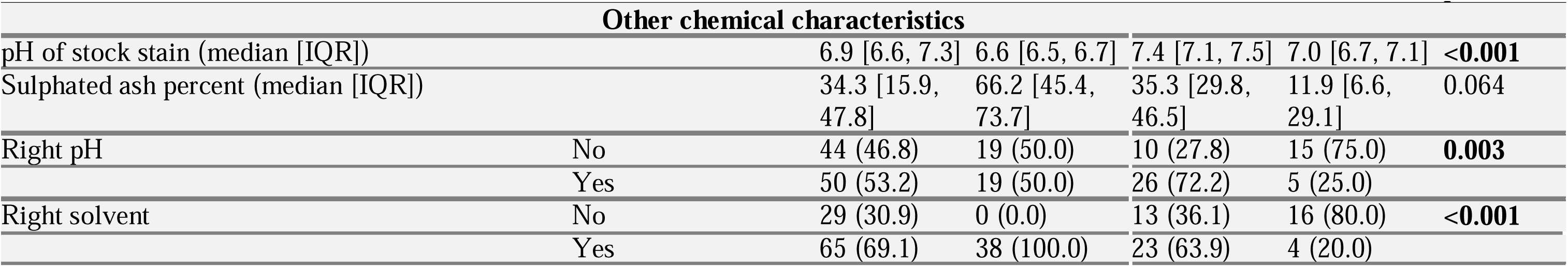
Physico-chemical profile of Rowmanowsky-type stains used for malaria microscopy in Plateau State, Nigeria stratified by stain type

### Ultraviolet Spectroscopic Characteristics of Stain Samples

The four most prominent groups of dyes present in the stain samples were detected in their order of relative abundance across stain samples and identified based on the maximum absorbance wavelengths (λmax) of the dye species present within each group. The first most conspicuous dyes had a median λmax of 624.0nm (IQR: 607.2, 658.0nm) for all stains, which corresponds to the λmax of Brilliant Cresyl Blue. When disaggregated by stain type, the median λmax was 658.5nm (IQR: 653.2, 660.0nm), corresponding to that of Methylene Blue for Field stain; 612.0nm (IQR: 517.0, 624.0nm), corresponding to Indigo carmine for the Giemsa stains; and 608.5nm (IQR: 516.8, 619.8nm), corresponding to Aniline Blue for the Leishman stains (p<0.001). Thus, Methylene Blue emerged as the most abundant dye accounting for 38.3% of dyes found in the first most prominent dyes in all stains but differ by stain type (Field stain: 81.6%; Giemsa stain: 13.9%; and Leishman stain: 0.0%, p<0.001). The highest prevalence of dye species irrelevant for the Romanowski-Giemsa effect (RGE) was found in the Leishman stains (Alcian Blue: 5.0%; Alizarin Red: 5.0%; Aniline Blue: 15.0%; Brilliant cresyl Blue: 5.0%; Sym-Dimethyl Thionine: 10.0%) and the Giemsa stains (Aniline Blue: 11.1%; Brilliant Cresyl Blue: 19.4%; Indigo Carmine: 11.1%; and Sym-Dimethyl Thionine: 2.8%). The second most prominent group of dyes also had a median λ_max_ of 624.0nm (IQR: 609.2, 624.0nm) for all stains, which corresponds to the λ_max_ of Brilliant Cresyl Blue. When disaggregated by stain type, the median λ_max_ was 624.0nm (IQR: 624.0, 624.0nm), corresponding to that of Brilliant Cresyl Blue for Field stain; 624.0nm (IQR: 590.8, 636.0nm), corresponding to Brilliant Cresyl Blue for the Giemsa stains; and 560.5nm (IQR: 516.8, 624.0nm), corresponding to Pyronine derivatives for the Leishman stains (p=0.004). Among the third most prominent group of dyes, the median λ_max_ was 539.0nm (IQR: 516.2, 600.5nm) for all stains, which corresponds to the λ_max_ of Neutral Red. The median λmax also differed by stain type. For the Field stains, median λ_max_ was 516.0nm (IQR: 515.0, 517.0nm), corresponding to the λ_max_ of Eosin B or Y; the median λ_max_ was 626.0nm (IQR: 544.2, 629.0nm) for the Giemsa stains, corresponding to the λ_max_ of Azure A; and among the Leishman stains, the median λmax was 549.0nm (IQR: 541.5, 599.5nm, p<0.001), which corresponds to the λmax of Phloxine B. It was also found that the presence of RGE-irrelevant dyes was more among the Leishman stains (Neutral red: 30.0%; Basic Fuchsin: 20.0%; Pyronine derivatives: 15.0%; and methylene Violet Bernthsen: 5.0%). Among the fourth most abundant dyes, the median λ_max_ was 542.0nm (IQR: 419.0, 624.0nm), corresponding to the λ_max_ of Neutral Red, which tend to differ across stain types (Field stain: 624.0nm [624.0, 624.0]nm; Giemsa stain: 517.5nm [410.0, 626.0nm]; and Leishman stain: 414.0nm [385.0, 543.5nm]; p<0.001). These groups of dyes were found to have a preponderance of unknown dyes (all stains: 36.2%; Field stain: 21.1%; Giemsa stain: 38.9%; Leishman stain: 60.0%). The most prevalent single dye detected in the stains were Methylene Blue in the Field stains (81.6%) and Eosin in the Giemsa stains (30.6%) and Leishman stains (35.0%). Azure b was detected as the most prevalent single dye in 7.4% of all stains (Field stain: 13.3%; Giemsa stain: 5.6%; and Leishman stain: 0.0%). The presence of RGE-antagonizing dyes was found in 8.5% of all stains (Field stain: 0.0%; Giemsa stain: 11.1%; and Leishman stain: 20.0%). (**Table 1**, **Fig 1-4**). The identities of the four most prominent dyes detected in each of the stain samples based on their λ_max_is shown Supplementary Tables 1-3 for Field, Giemsa, and Leishman stains respectively.

**Fig 1.**
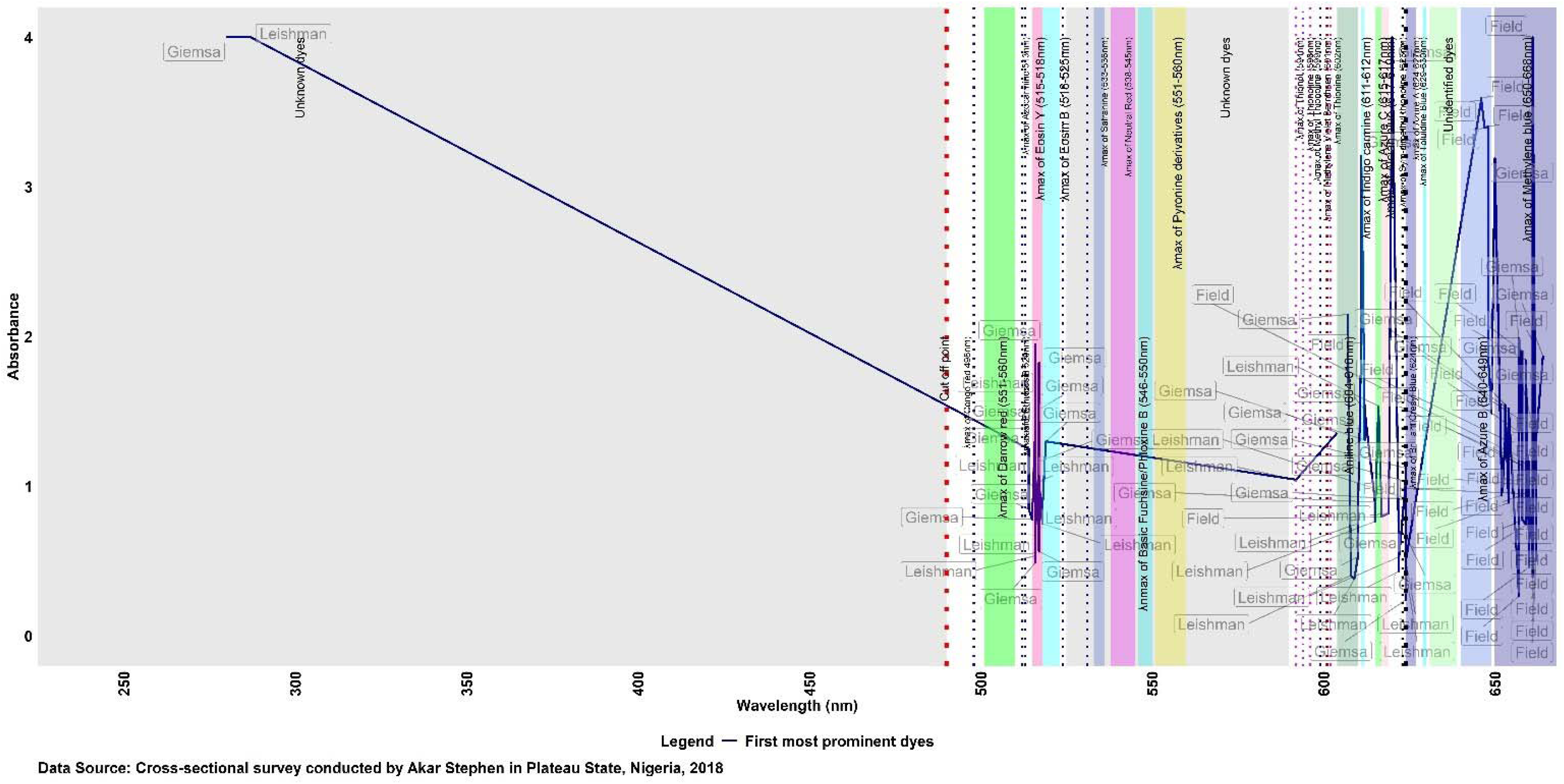
Spectroscopic characteristics of the first prominent group of dyes present in Romanowski-type stains used for malaria microscopy in Plateau state, Nigeria disaggregated by mode of preparation”,

**Fig 2.**
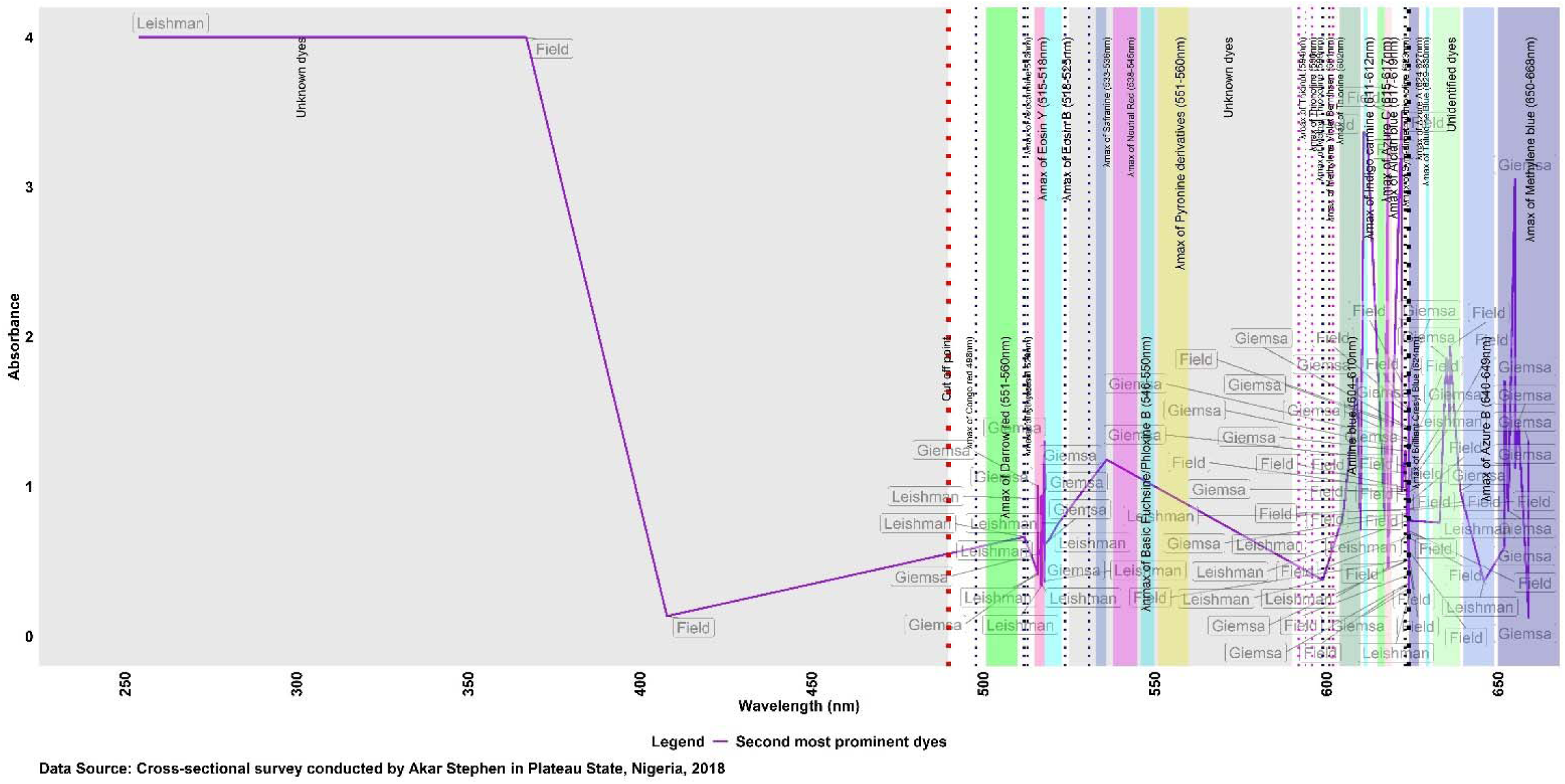
Spectroscopic characteristics of the second most prominent dyes present in Romanowski-type stains used for malaria microscopy in Plateau state, Nigeria disaggregated by stain type”,

**Fig 3.**
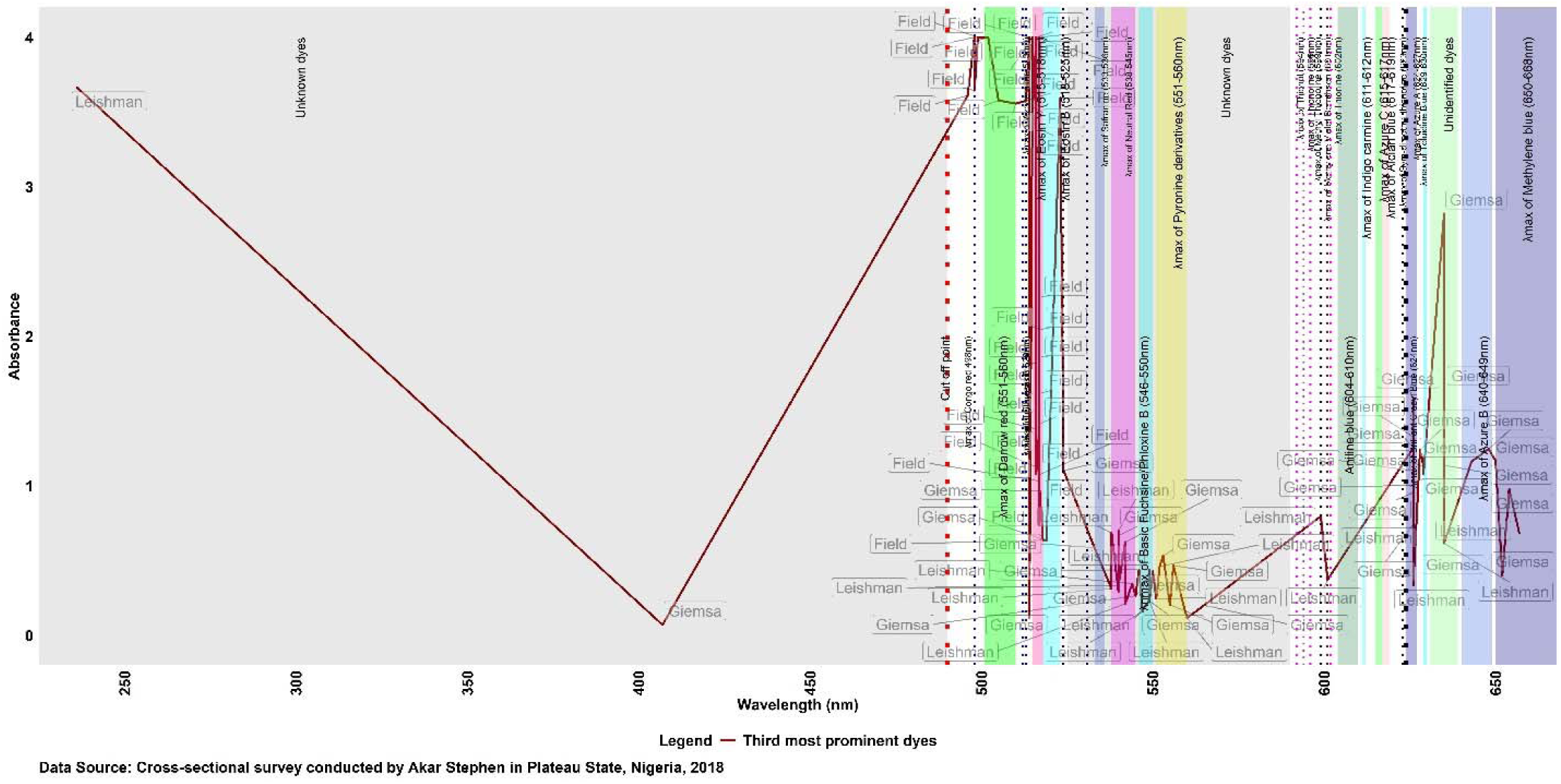
Spectroscopic characteristics of the third most prominent group of dyes present in Romanowsky-type stains used for malaria microscopy in Plateau state, Nigeria disaggregated by stain type.

**Fig 4.**
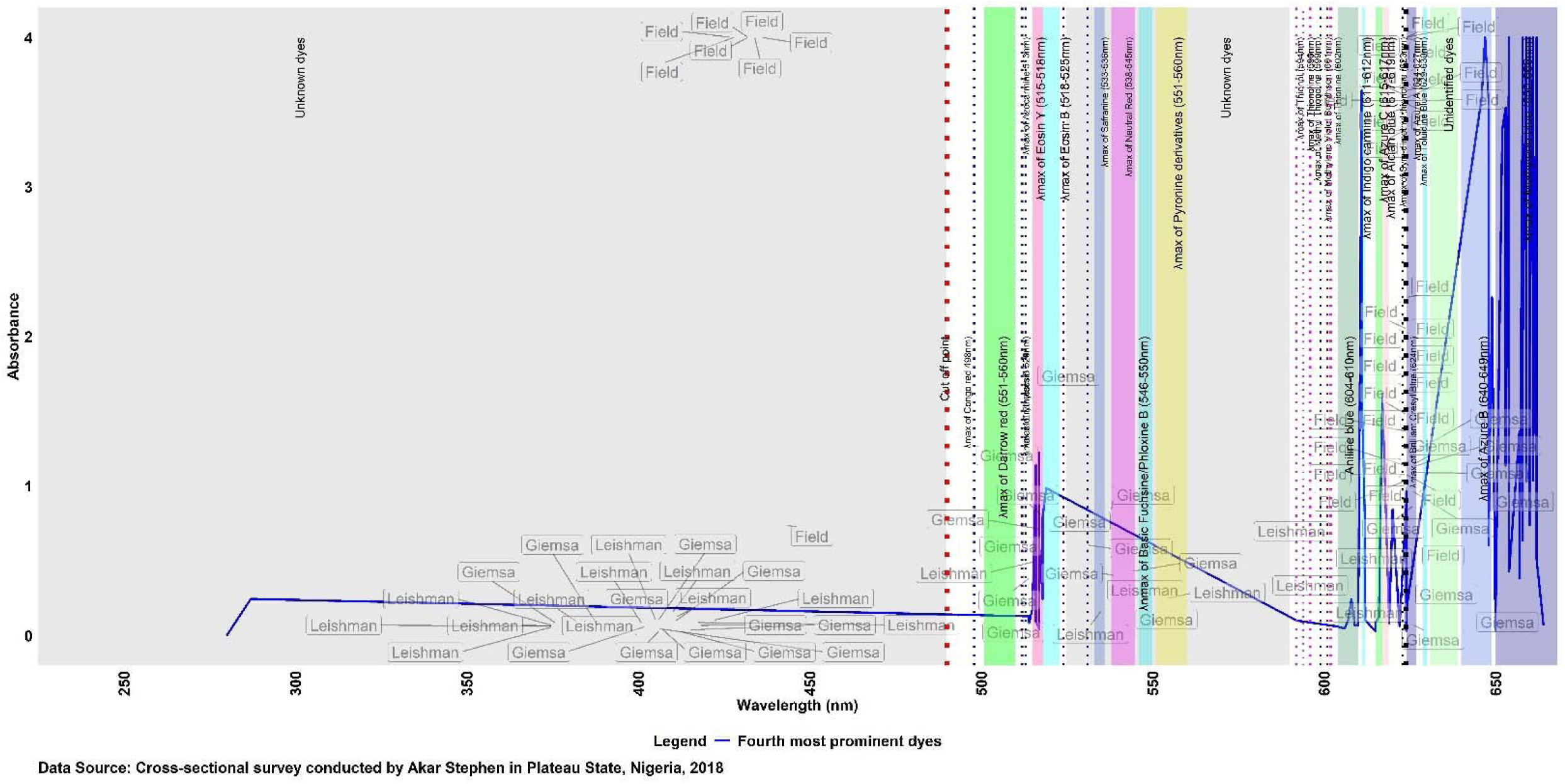
Spectroscopic characteristics of fourth most prominent group of dye present in Romanowsky-type stains used for malaria microscopy in Plateau state, Nigeria disaggregated by stain type.

### Determination of Dye Proportions by High Performance Liquid Chromatography (HPLC)

HPLC was done to determine the proportion of the dyes essential for the Romanowsky-Giemsa effect (RGE) in each of the stain samples. The dyes analyzed were pure methylene blue, Azure A, Azure B, eosin B and eosin Y. The presence and concentrations of these dyes were determined using the retention times of the standard dyes. Individual solutions of methylene blue, Azure A, Azure B, eosin B and eosin Y prepared from pure powders were chromatographed in triplicates by HPLC to determine the individual mean retention time of the eluents. The retention times were used to identify the presence of the dyes in the samples of stains collected. The retention time obtained for each dye species were as follows: Eosin Y, 1.4 minutes, Eosin B, 1.404 minutes, methylene blue, 2.920 minutes and Azure B, 2.923 minutes. The dye peak area under the HPLC graph output, which was calculated as the area of a triangle, was used to estimate the proportion of each dye species present in the stain samples.

The median proportion of Azure A for all stains was found to be 14.6% (IQR: 10.5-23.9%), which in turn differ significantly by stain type (Field stain: median = 12.6%, IQR = 10.6-14.6%; Giemsa stain: median = 23.2%, IQR = 14.0-29.9%; and Leishman stain: median = 16.3%; IQR = 5.4-26.0%, p=0.001). The median proportion of Azure B for all stains was 9.2% (IQR: 0.0-15.2%) and vary significantly by stain type (Field stain: median = 12.9%, IQR = 6.6-14.8%); Giemsa stain: median = 12.5%, IQR= 4.5-22.2%; and Leishman stain: median = 0.0, IQR= 0.0-0.0%, p<0.001). For Eosin B, median proportion for all stains was 0.8% (IQR: 0.0-6.6%), which was found to differ across stain types (Field stain: median = 0.0%, IQR = 0.0-0.6%); Giemsa stain: median = 3.2%, IQR= 0.0-6.6%; and Leishman stain: median = 6.1, IQR= 0.7-7.9%, p<0.001). The median proportion of Eosin Y was found to be 7.6% (IQR: 2.6-21.4%) which also differ significantly across stain types (Field stain: median = 2.2%, IQR = 1.2-4.0%); Giemsa stain: median = 10.7%, IQR= 6.9-15.7%; and Leishman stain: median = 27.5, IQR= 23.9-28.8%, p<0.001). For Methylene Blue, the median proportion was found to be 2.9% (IQR: 0.0-30.5%) for all stains, which was found to differ by stain type (Field stain: median = 32.5%, IQR = 23.2-38.1%); Giemsa stain: median = 1.6%, IQR= 0.0-2.6%; and Leishman stain: median = 0.0, IQR= 0.0-0.0%, p<0.001). The proportion of unidentified dyes (unknown dyes) was found not to vary significantly across tains types (All stains: medina =39.5% [IQR: 33.7, 50.1%]; Field stain: median = 37.8% [IQR: 33.5, 46.3%]; Giemsa stain: median = 41.0% [IQR: 33.8, 46.3%]; and Leishman stain: median = 45.5% [IQR: 34.3, 66.5%], p=0.343. However, the proportion of RGE-relevant dyes differ significantly across stain types (All stains: median = 35.4% [IQR: 25.4, 48.6%]; Field stain: median = 50.5% [IQR: 37.3, 55.9%]; Giemsa stain: median = 29.0% [20.7, 39.3%]; and Leishman stain: median = 27.9% [24.7, 30.1%], p<0.001). The ratio of Azure b to Eosin Y, which are the crucial ingredients of the RGE reaction were also found to differ significantly across the stain types (All stain: median = 0.1 [IQR: 0.0, 2.6]; Field stain: 2.7 [0.4, 3.2]; Giemsa stain: 0.1 [0.0, 0.2]; and Leishman stain: 0.0 [0.0, 0.0], p<0.001) (**Table 1**). On comparing the dye proportions by mode of stain preparation, it was found that commercially prepared Giemsa stain solution had higher proportion of Azure A compared to in-house solutions (**Fig 5a**). The proportion of Azure B was found to be lowest among commercially prepared Leishman stain solutions and highest in in-house Giemsa stain solutions (**Fig 5b**). Both commercial and in-house Field stain solutions contained higher proportion of Methylene Blue compared to Giemsa and Leishman stains (**Fig 5c**). The Field stains contained lesser proportion of Eosin B compared to Giemsa and Leishman stains irrespective of mode of stain preparation (**Fig 5d**). Both commercial and in-house solutions of Leishman stains were found to contain more Eosin Y compared to Field and Giemsa stains (**Fig 5e**), but the Leishman stains exhibited the least Azure B to Eosin Y ratio while the Field stains were highest (**Fig 5f**). The proportion of Azure A, Azure B, Eosin B, Eosin Y, and Methylene Blue present in the individual stain samples are shown in **Supplementary Tables 3-4** for Field, Giemsa, and Leishman stains respectively.

**Fig 5.**
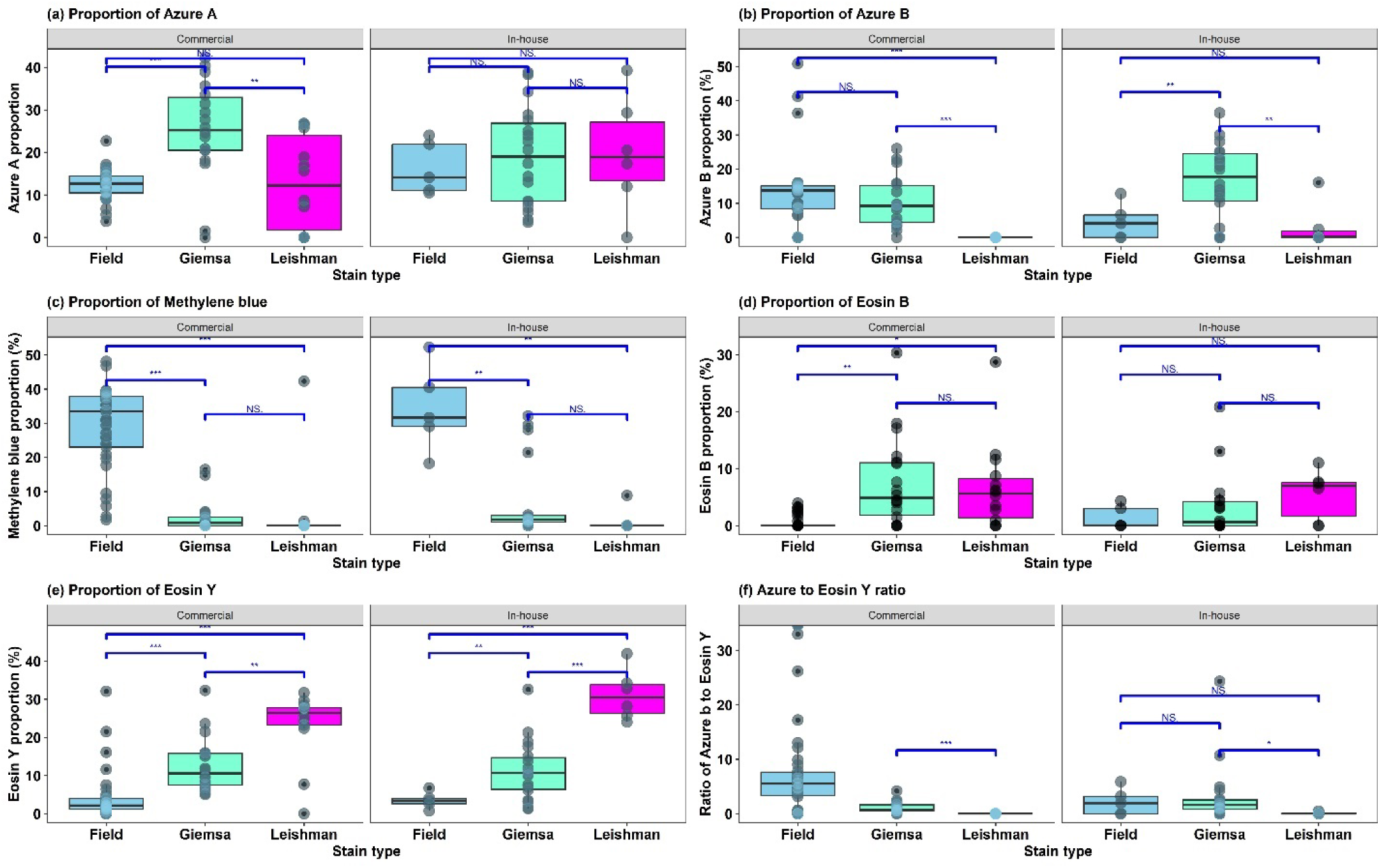
Comparison of the proportion Azure A, Azure B, Methylene Blue, Eosin B, Eosin Y and the ratio of Azure B to Eosin Y present in Field, Giemsa, and Leishman stains used for malaria microscopy in Plateau State, Nigeria.

### Stain pH, Sulphated Ash Percent and Solvent Used for Preparation Stain solution

Median pH of stock stain solution was 6.9 (IQR = 6.6 – 7.3) for all stains (Field stains: median = 6.6 [IQR: 6.5 – 6.7]; Giemsa stains: median = 7.4 [IQR = 7.1–7.5]; and Leishman stains: median = 7.0[6.7-7.1], p<0.001). The median sulphated ash percent of dried powders from which in-house stain solutions were made was 34.3% (15.9– 47.8%) for all stains (Field stain: median = 66.2% [IQR = 45.4– 73.7%]; Giemsa stain: median = 35.3% [29.8-46.5%]; and Leishman stain: median = 11.9% [IQR = 6.6 – 29.1%], p=0.064) (**Table 1**). All Field stain solutions were found to be made of water as solvent. Only 20% (4/20) of Leishman stains were found to be made of absolute methanol. Up to 63.9% (23/36) of Giemsa stains were found to be composed of a glycerol to absolute methanol ratio of 1:1 with a median of 1.3 (0.9 – 1.9). Glycerol to methanol ratio was found to be higher in in commercially prepared stain solution (median = 1.8; IQR: 0.9-2.7) compared to in-house stain solution (median = 1.2; IQR: 1.1-1.5) of Giemsa stains (**Fig 6a**).

**Fig 6.**
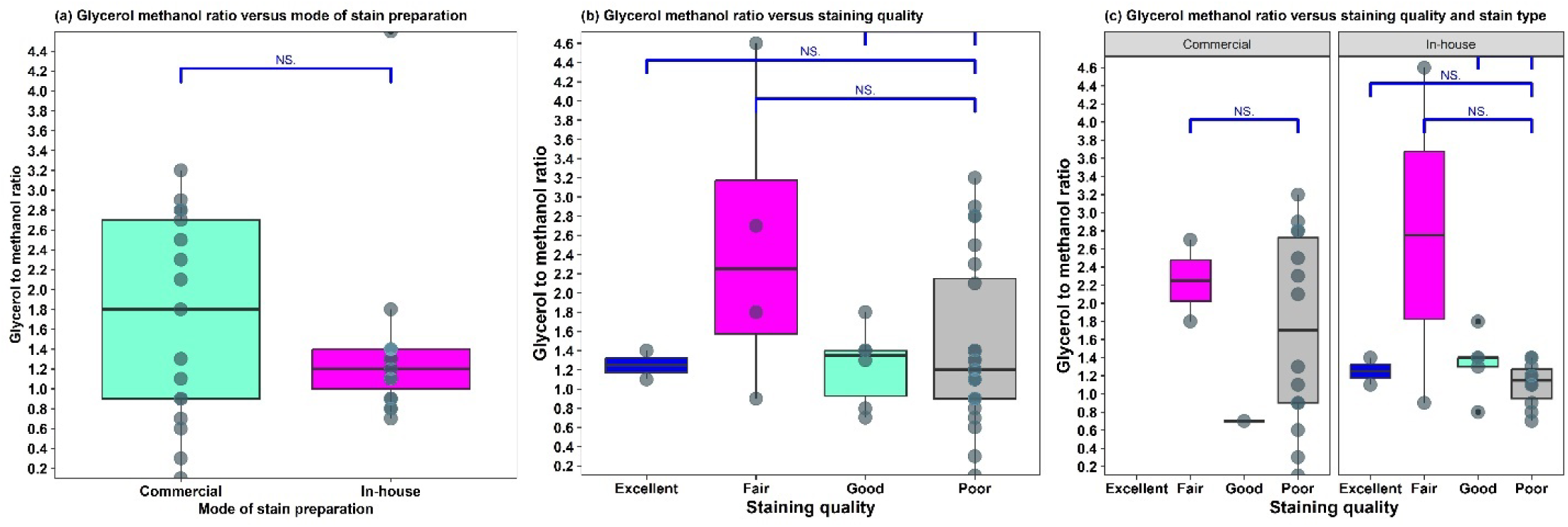
Comparison of Glycerol to methanol ratio of Giemsa stains used for malaria microscopy in Plateau State, Nigeria.

### Assessment of Staining Quality of Stain Samples

The staining quality of the stain samples was assessed using thin blood film microscopy. Known malaria positive blood smears were stained and examined by WHO certified malaria microscopists. Staining quality of each stain sample was measured by the quality of colour imparted by the stain samples on the cytoplasm and nucleus of malaria parasites. Staining quality was graded excellent if the cytoplasm was stained blue and nucleus pink; good if cytoplasm was stained blue and nucleus red or pale pink; fair if cytoplasm was stained purple-blue and nucleus deep red or pale pink; and poor if cytoplasm was pale blue, purple or unstained and nucleus. Of the two stain samples that yielded excellent staining, one was a the reference Giemsa stain sample obtained from ANDi laboratory, Lagos and the other a Giemsa-type stain described by Marshall et al. (1975). Among stains that were graded as “good”, 75.0% were Giemsa and 25.0% were Field stains. None of the Leishman stains yield “good” staining. Stains that yielded “fair” (81.2%) or “poor” (70.6%, p=0.027) staining outcome were largely commercially prepared solutions, which were also had expiry dates missing from the labels on the containers (75.0% and 57.4% respectively). “Poor” performing stains were found to contain RGE-irrelevant dyes such as Alcian Blue, Alizarin Red, Aniline Blue, Azure C, Brilliant Cresyl Blue, Indigo Carmine, Methylene Blue, Sym-Dimethyl Thionine, Methyl Thionine, Safranine, Basic Fuchsin, Congo Red, Darrow Red, Erythrosin B, Methylene Violet Bernthsen, Neutral Red, Pyronine derivatives, Toluidine Blue, and unidentified (unknown) dyes (**Table 2**).

**Table 2.**
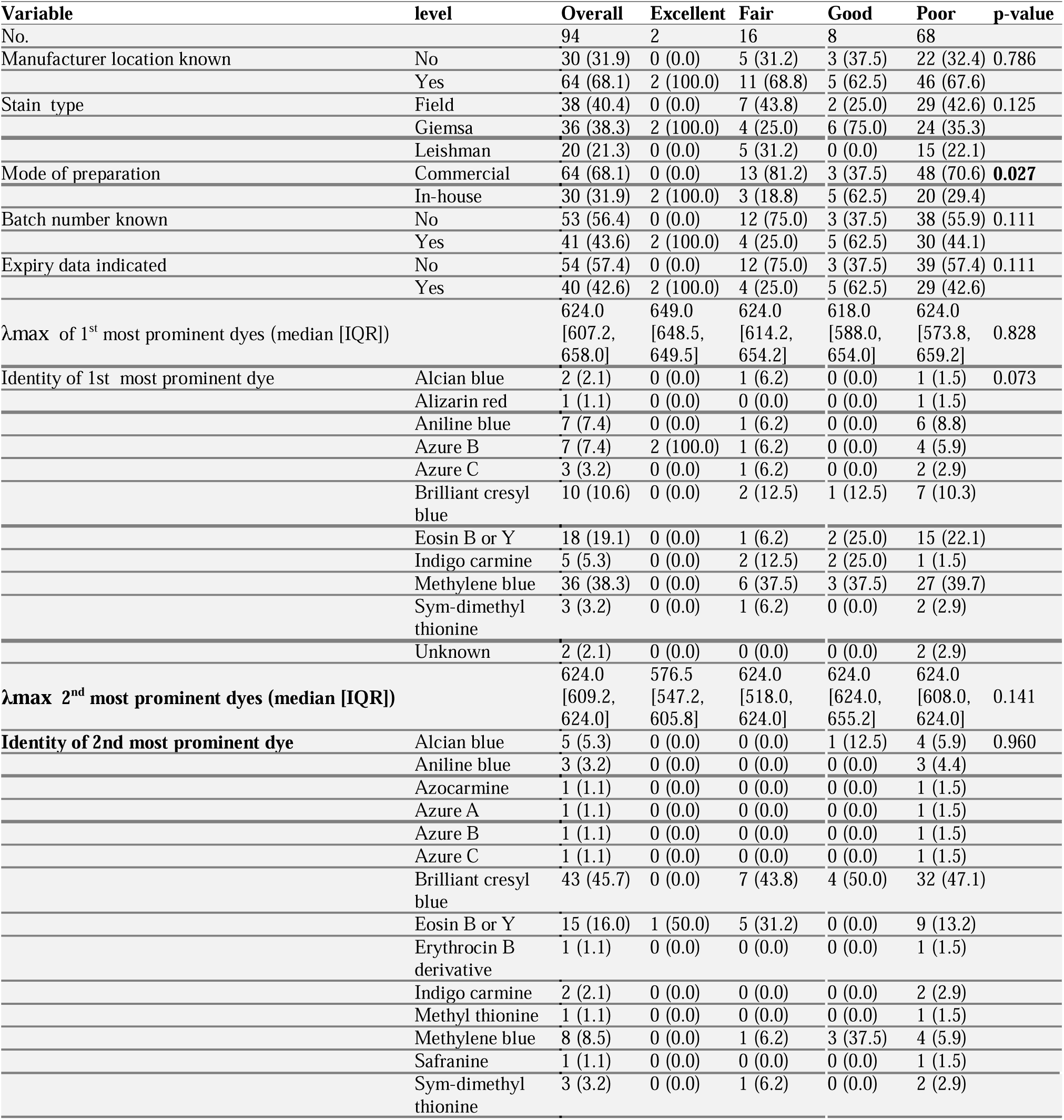
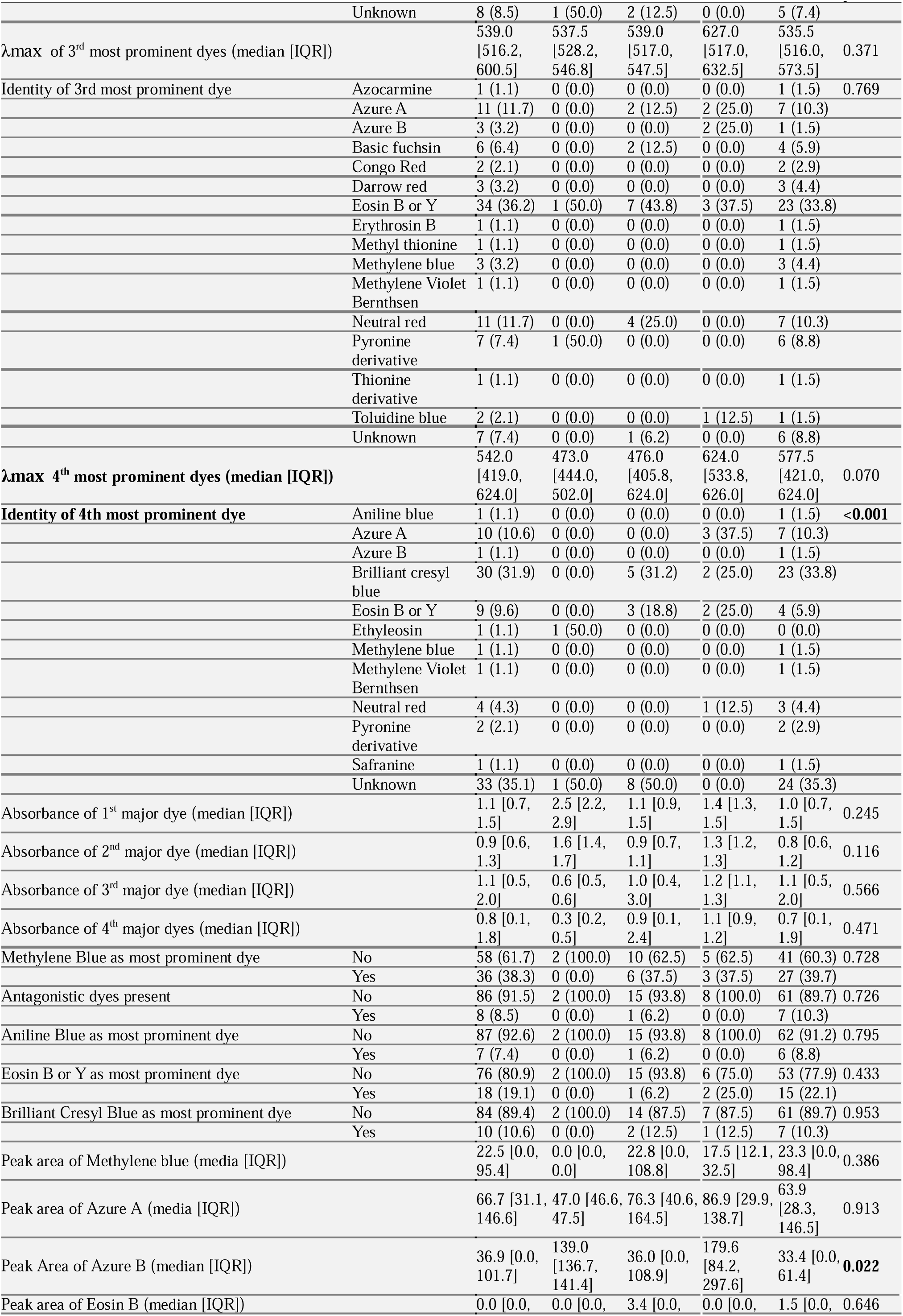
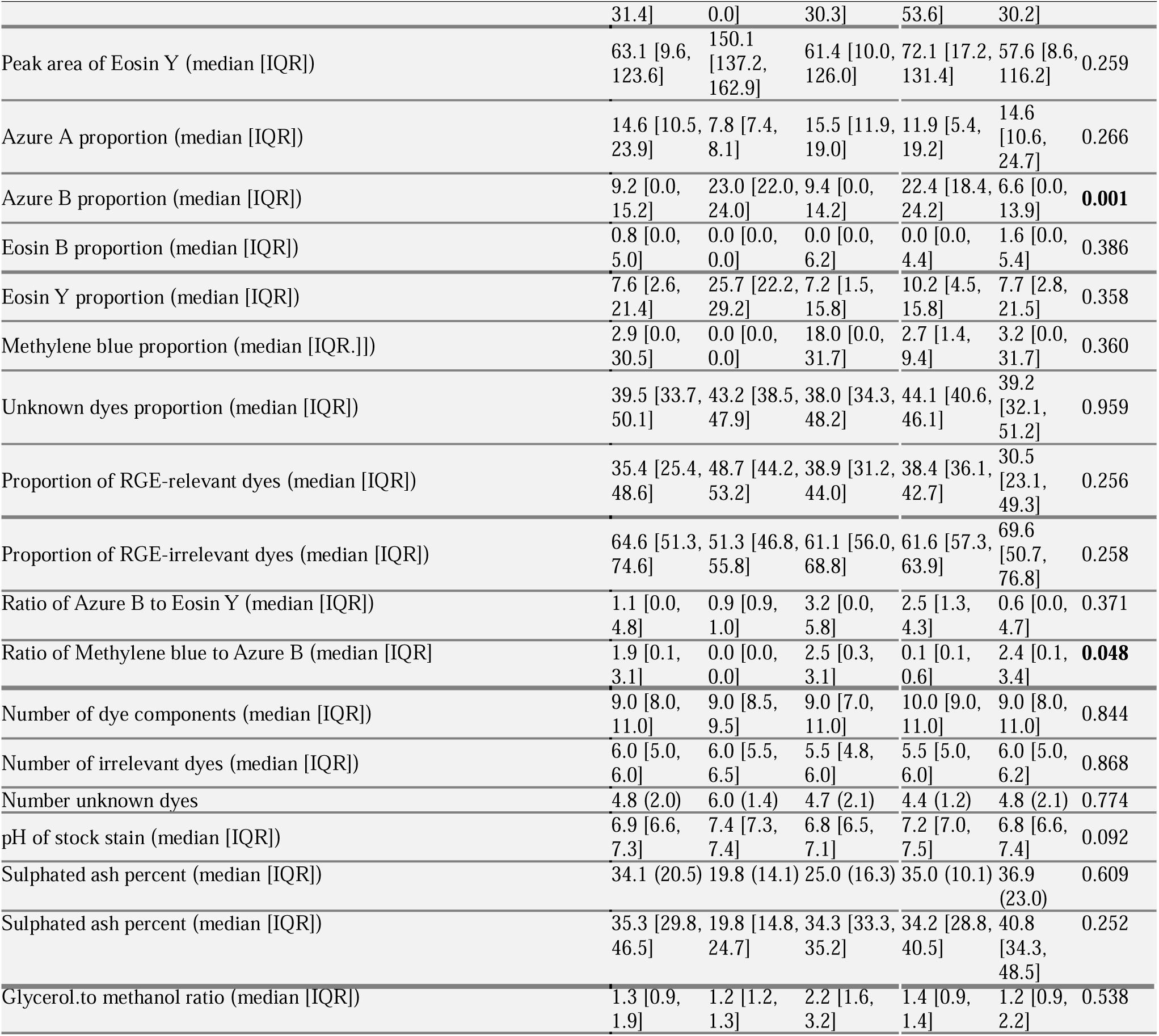
Evaluation of the performance of Romanowsky-type stains used for malaria microscopy in Plateau State, Nigeria.

The distribution of the maximum absorbance wavelengths of the four most prominent group of dyes by the staining performance is shown in supplementary figures 1-4. They also had a higher presence of known RGE antagonistic dyes (10.3%). “Poor” stains were found to contain higher proportion of Azure A (median = 14.6%; IQR: 10.6-24.7%); significantly lower proportion of Azure B (median = 6.6%; IQR: 0.0-13.9%, p=0.001); higher proportion of Eosin B (median = 1.6%; IQR: 0.0-5.4%); lower Eosin Y (median = 7.7%; IQR: 2.8-21.5%); lower proportion of RGE-relevant dyes (median = 30.5%; IQR: 23.1-49.3%); higher Methylene Blue to Azure B ratio (median = 2.4; IQR: 0.1-3.4, p=0.048); and higher sulphated ash percent (median = 40.8%; IQR: 34.3-48.5%) (Table 2). A comparison of the proportion of the Azure, Azure B, Eosin B, Eosin Y, and Methylene Blue present in the stains by staining outcome and stain type is shown in **Fig 7** while **Fig 8** shows that for Azure B to Eosin Y ratio, Methylene Blue to Azure B ratio, proportion of RGE-relevant dyes, and proportion of RGE-irrelevant dyes. “Good” performing Field stains had a median stock pH of 6.8 while “poor” stains exhibited a median stock pH of 6.6. “Excellent” and “good” performing Giemsa stains had a median stock pH of about 7.2 with “good” performing stains exhibiting a much wider inter-quartile range. For the Leishman stains, the “poor” performing stain samples exhibited a higher pH compared to those that performed “fairly” (**Fig 9a and b**). It was found that the higher the sulphated ash percent of the stain powder the poorer the performance of the stain solution, with the Field stains exhibiting the highest sulphated ash percent (**Fig 9c and d**). For the Giemsa stains, “excellent” and “good” performing stains had a much narrower interquartile range. Variation in glycerol to methanol ratio was found be wider among commercially prepared Giemsa stain solutions (**Fig 6b and c**).

**Fig 7.**
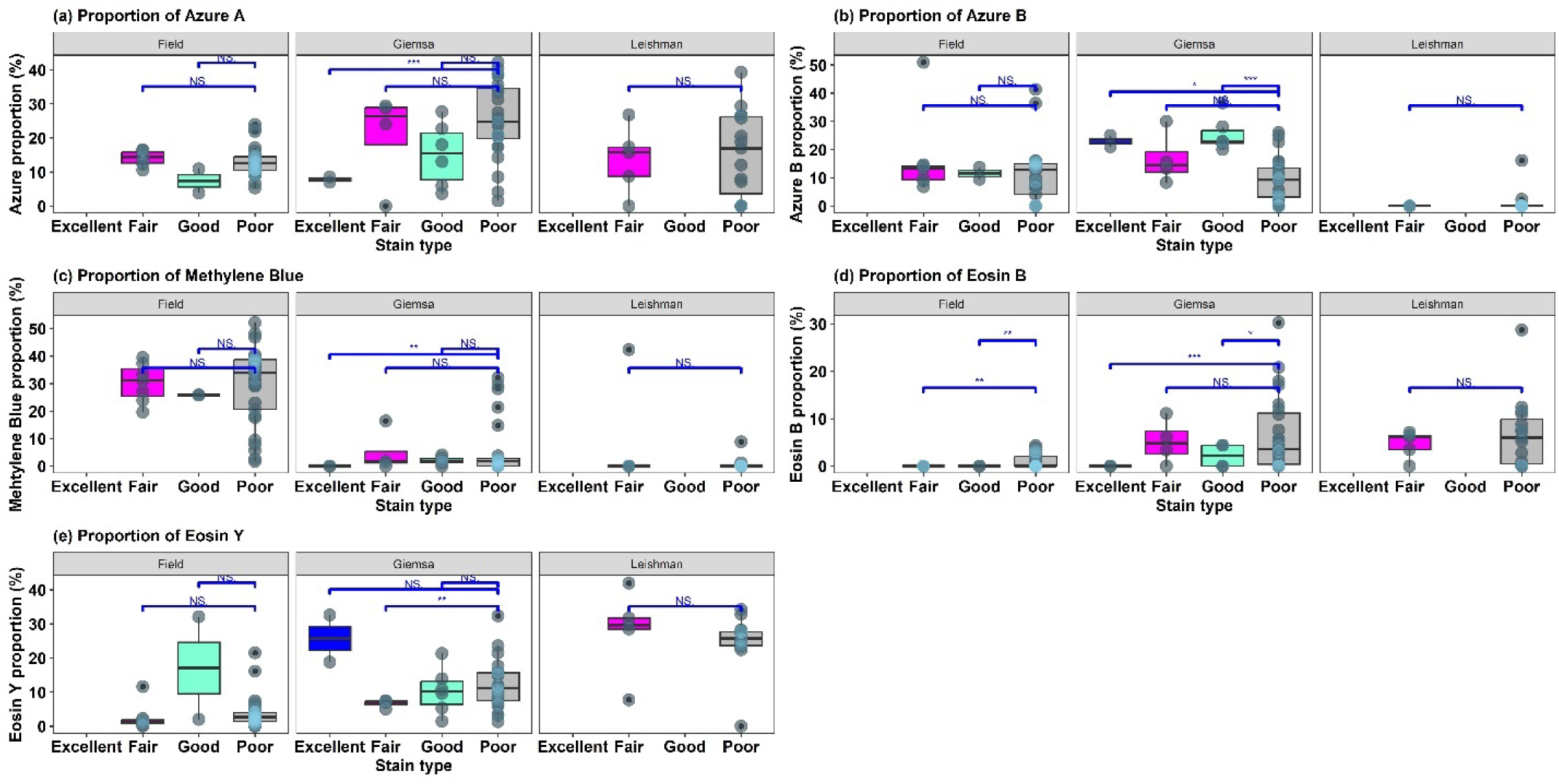
Comparison of proportion of Azure A, Azure B, Methylene Blue, Eosin B, and Eosin Y present in Romanowsky-type stains used for malaria microscopy in Plateau State by staining quality.

**Fig 8.**
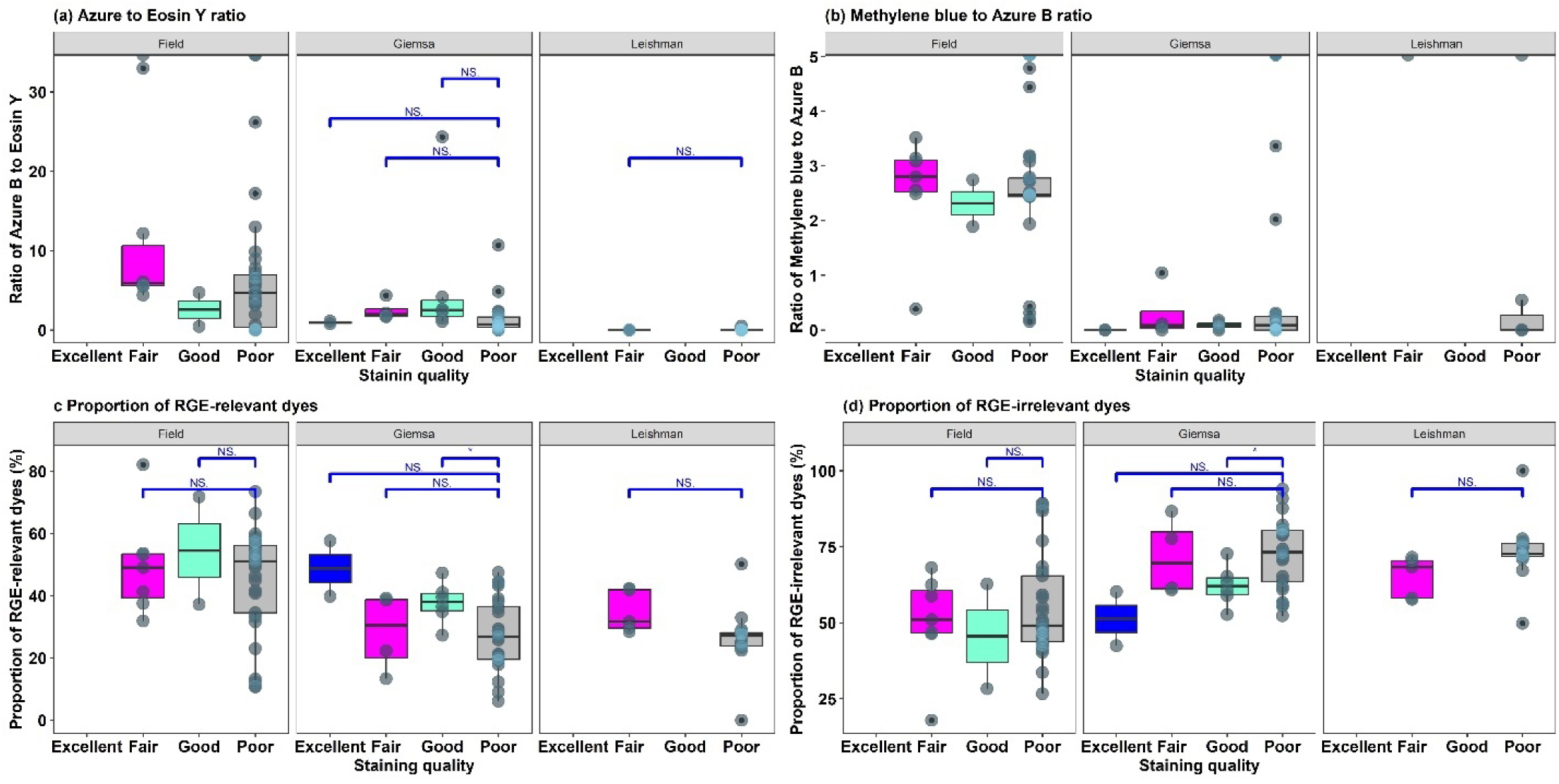
Comparison of proportion of Azure B to Eosin Y ratio, Methylene Blue to Azure B ratio, Proportion of RGE-relevant dyes, and Proportion of RGE-irrelevant dyes present in Romanowsky-type stains used for malaria microscopy in Plateau State by staining quality.

**Fig 9.**
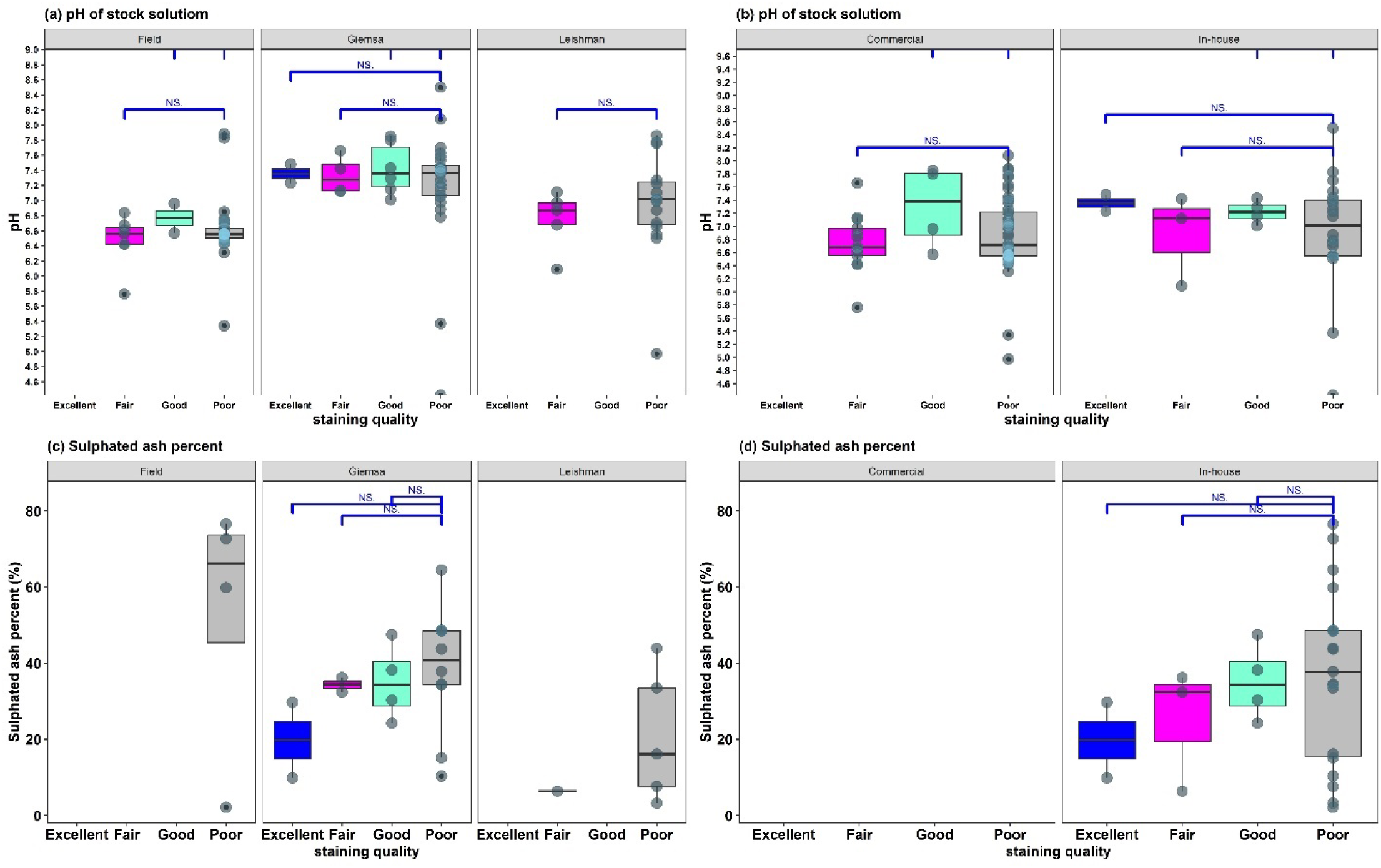
Comparison of pH of stock solution of Romanowsky-type stains used for malaria microscopy in Plateau State, Nigeria by stain type and staining quality.

### Factors Associated with Quality of Staining

In determining the factors associated with satisfactory staining outcome (quality of staining), we combined stains with “fair” and “good” staining outcome as “satisfactory” (coded as 1) and those with “poor” staining outcome as “unsatisfactory” (coded as 0). At bivariate analysis, it was found that having a proportion of Azure B greater than 20% was the only factor associated with satisfactory staining outcome (crude OR = 7.88, 95% CI: 2.45-28.40, p<0.001). However, stain type being Giemsa (crude OR = 1.61, 95% CI: 0.58-4.57, p=0.40); in-house prepared stains (crude OR = 1.5, 95% CI: 0.57-3.58, =0.40); alcohol-based stains (crude OR= 1.40, 95% CI: 0.56-3.71, 0.50); most prominent dye being Azure B (crude OR= 2.09, 95% CI: 0.39-10.20, p=0.40); first four most prominent dyes inclusive of RGE-relevant dyes (crude OR= 1.64, 95% CI: 0.32-7.25, p= 0.50); proportion of Azure A less than 20% (crude OR= 2.20, 95% CI: 0.82-6.64, p=0.14); proportion of Eosin B less than 5% (crude OR= 1.63, 95% CI: 0.57-5.42, p=0.40); Azure B to Eosin Y ratio greater than 1.5 (crude OR= 2.23, 95% CI: 0.89-5.79, p=0.09); Methylene Blue to Azure B ratio less than 1 (crude OR= 1.54, 95% CI: 0.57-4.20, p= 0.40); pH of stock stain solution (crude OR= 1.23; 0.60-2.67, p=0.60), and sulphated ash percent of less than 30% (crude OR= 1.92, 95% CI: 0.45-7.57, p-value= 0.4) were associated with satisfactory staining outcome but not statistically significant. Independent predictors of satisfactory staining outcome were proportion of Azure B greater than 20% (adjusted OR= 15.10, 95% CI: 2.62-115.00, p= 0.007); Azure B to Eosin Y ratio greater than or equal to 1.5 (aOR= 4.55; 95% CI: 1.03-25.1, p=0.004); and stains solution formulated with an alcohol-based solvent (adjusted OR= 27.70, 95% CI: 2.43-462.00, p= 0.012) (**Table 3**). Similar predictors were obtained when the analysis was stratified by mode of stain preparation (**Table 4**).

**Table 3.**
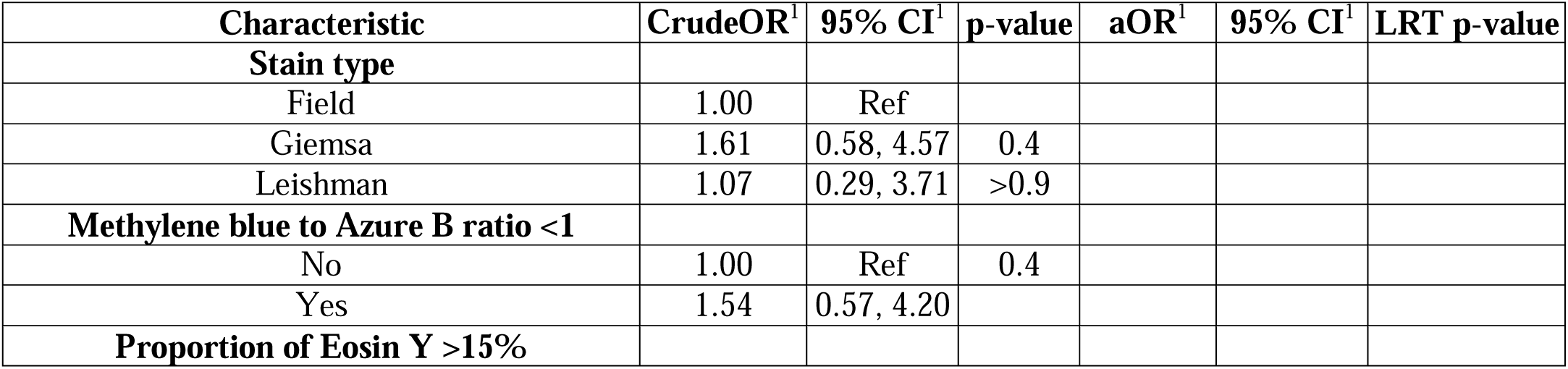
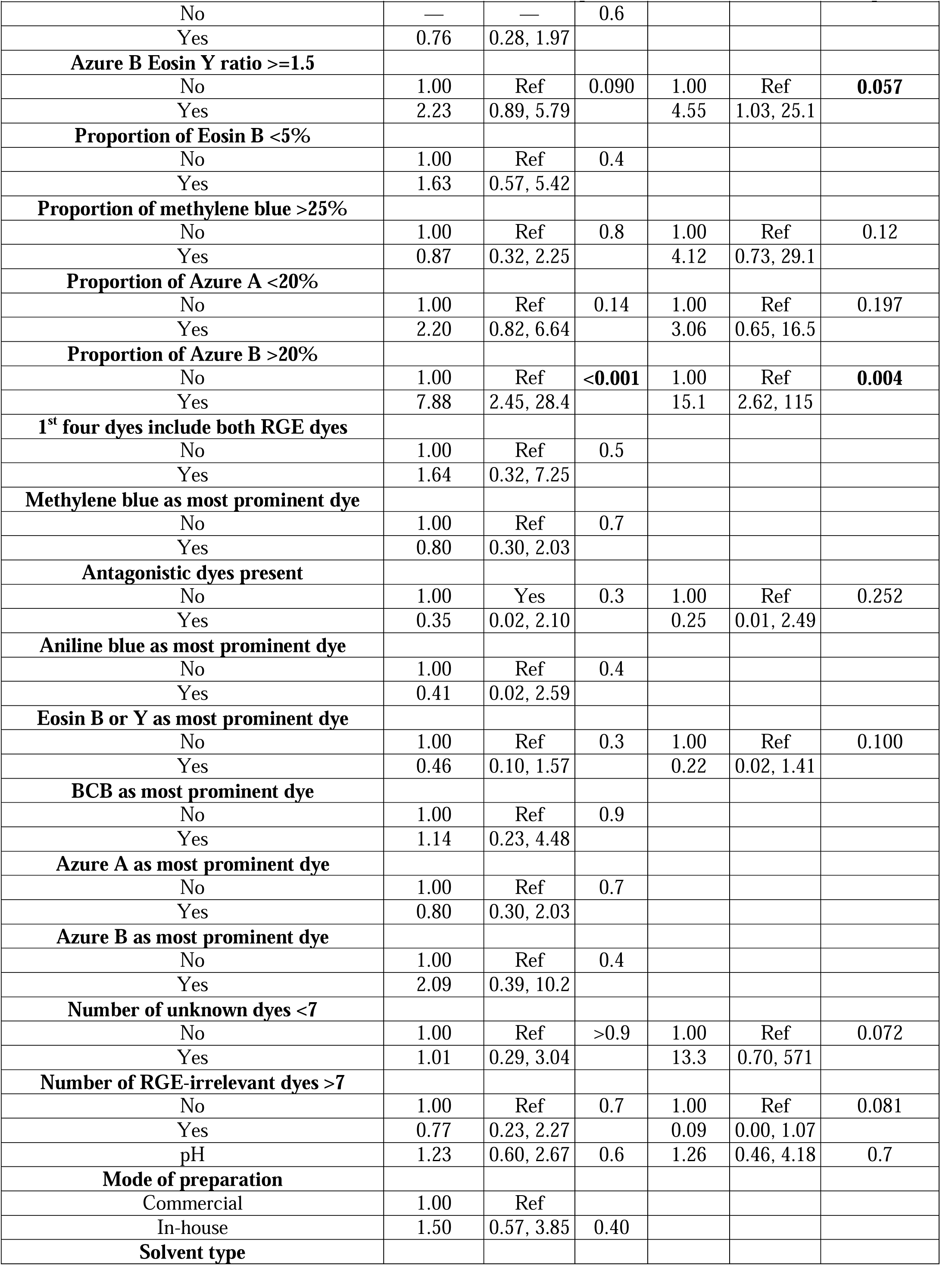
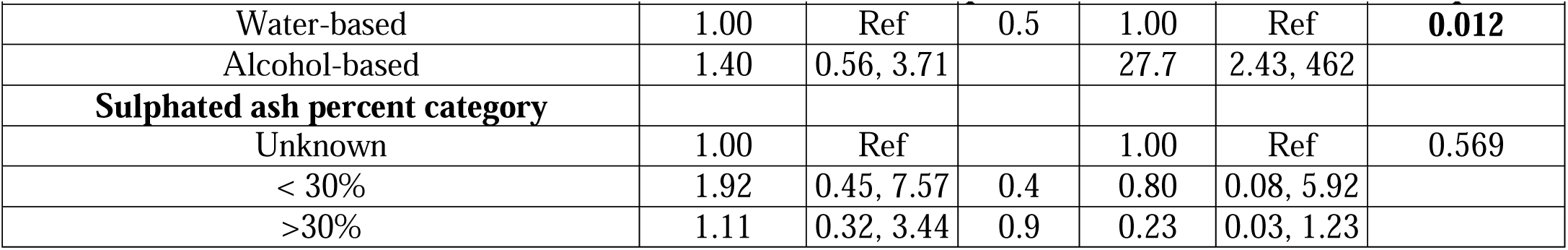
Factors associated with satisfactory staining outcome among Romanowsky-type stains used for malaria microscopy in Plateau State, Nigeria.

**Table 4.** Predictors of satisfactory staining outcome among Romanowsky-type stains used for malaria microscopy in Plateau State, Nigeria stratified by mode of stain preparation

| Characteristic | Commercially Prepared Stains |  |  | In-house Prepared Stains |  |  |
| --- | --- | --- | --- | --- | --- | --- |
|  | OR<sup>1</sup> | 95% CI<sup>1</sup> | p-value | OR<sup>1</sup> | 95% CI<sup>1</sup> | p-value |
| Azure B to Eosin Y ratio >=1.5 |  |  |  |  |  |  |
| No | 1.00 | Ref | 0.2 | 1.00 | Ref | 0.2 |
| Yes | 3.40 | 0.84, 15.8 | 0.095 | 4.55 | 1.03, 25.1 | 0.057 |
| Proportion of Azure A <20 |  |  |  |  |  |  |
| No | 1.00 | Ref | 0.12 | 1.00 | Ref | 0.2 |
| Yes | 3.30 | 0.76, 16.6 |  | 3.06 | 0.65, 16.5 |  |
| Proportion of Methylene blue >2 |  |  |  |  |  |  |
| No | 1.00 | Ref | 0.2 | 1.00 | Ref | 0.12 |
| Yes | 3.07 | 0.59, 19.1 |  | 4.12 | 0.73, 29.1 |  |
| Presence of antagonistic dyes |  |  |  |  |  |  |
| No | 1.00 | Ref | 0.3 | 1.00 | Ref | 0.3 |
| Yes | 0.27 | 0.01, 2.62 |  | 0.25 | 0.01, 2.49 |  |
| Eosin B or Y as most prominent dye |  |  |  |  |  |  |
| No | 1.00 | Ref | 0.3 | 1.00 | Ref | 0.15 |
| Yes | 0.35 | 0.04, 1.95 |  | 0.22 | 0.02, 1.41 |  |
| Proportion of Azure B >20% |  |  |  |  |  |  |
| No | 1.00 | Ref | **0.006** | 1.00 | Ref | **0.004** |
| Yes | 11.4 | 2.24, 78.5 |  | 15.1 | 2.62, 115 |  |
| Number of unknown dyes >7% |  |  |  |  |  |  |
| No | 1.00 | Ref | 0.076 | 1.00 | Ref | 0.11 |
| Yes | 16.3 | 1.01, 625 |  | 13.3 | 0.70, 571 |  |
| Number of irrelevant dyes >7% |  |  |  |  |  |  |
| No | 1.00 | Ref | **0.049** | 1.00 | Ref | 0.081 |
| Yes | 0.07 | 0.00, 0.75 |  | 0.09 | 0.00, 1.07 |  |
| pH | 1.19 | 0.47, 3.50 | 0.7 | 1.26 | 0.46, 4.18 | 0.7 |
| Solvent type |  |  |  |  |  |  |
| Water-based | 1.00 | Ref | **0.035** | 1.00 | Ref | **0.018** |
| Alcohol-based | 13.3 | 1.51, 158 | **0.028** | 27.7 | 2.43, 462 | **0.012** |
| Sulphated ash percent category |  |  |  |  |  |  |
| Unknown |  |  |  | 1.00 | Ref | 0.11 |
| <30% |  |  |  | 0.80 | 0.08, 5.92 |  |
| >30% |  |  |  | 0.23 | 0.03, 1.23 |  |

<sup>1</sup>OR = Odds Ratio, CI = Confidence Interval

## Discussion

For the first time, to our knowledge, an attempt has been made to profile the physico-chemical characteristics of the Romannowsky-type (Rowmanowsky-Giemsa stains) used for malaria microscopy in a malaria endemic setting in sub-Saharan Africa. We found that laboratories utilize Romanowsky-type stains other than the WHO recommended Giemsa stain for malaria microscopy. This may probably indicate lack of access to or non-adherence to the WHO malaria microscopy. However, this finding is similar to that reported in a study in India in which preference is given to the use of Leishman rather than Giemsa stain (Sathpathi et al., 2014). In about half of stain samples, it was found that batch number and expiration dates were missing from the product label and some manufacturers failed to indicate their company address. None of the manufacturers listed the dye components of their stains on the container label of the stains. Issues of proper labelling of commercially available Romanowsky-type stains have been reported previously (Marshall, 1974; Schulte, 1991). To achieve certification, the Biological Stains Commission (BSC) requires stain manufacturers to these critical details on their product labels (Penney et al., 2002; Schulte, 1991). Most manufacturers were found to have demonstrated evidence of non-compliance with these requirements in this study. These findings highlight the need for adequate regulation of biological stains used for clinical laboratory diagnostics in Nigeria. Ultraviolet spectroscopic and HPLC assays have revealed a high degree of variability of dye composition and the presence of RGE-irrelevant and RGE-antagonizing dyes in the stain samples analyzed in this study. Whereas the two most prominent dyes in our reference stain (Marshall et al., 1975c) were Azure B and Eosin Y, which are actually the only two dye species needed to produce the Romanowsky-Giemsa Effect (Friedrich and Seiffert, 1990; Horobin, 2011; Wittekind, 2008; Wittekind and Gehring, 1985), majority of the stain samples from various laboratories exhibited a wide range of variability with regards to the presence and proportion of these two essential dyes. This finding is similar to the results of previous commercial stains evaluation (Proctor and Horobin, 1985; Horobin and Walter, 1987a; Marshall et al., 1975a; Schulte, 1991). Such variability in dye composition and concentrations has been incriminated in capricious and poor performance of Romanowsky-type stains (Marshall, 1978a; S. A. Bentley, 1980; Schulte, 1991). Variation in the proportion of the RGE-relevant dyes was conspicuous even amongst stains produced by the same manufacturer. This is not surprising as previous studies have demonstrated evidence of batch-to-batch variation in the dye composition of commercial Romanowsky-type stains (Lubrano et al., 1977; Marshall et al., 1978, 1975c; Marshall, 1976b). It has been reported that large-scale manufacture of Romanowsky-Giemsa stains rely heavily on the polychroming of Methylene blue (Krafts et al., 2010; Lillie, 1942), which has been shown to be more difficult to control at industrial scale, resulting in the formation various RGE-irrelevant dyes in addition to Azure B (Horobin, 2011; Lillie, 1942; Löhr et al., 1974; Marshall, 1976b). This may have been responsible for the presence of various spurious dyes detected in the stain samples, such as Aniline Blue and Brilliant Cresyl Blue. We identified several RGE-irrelevant dye species present in the stain solutions based on their uv-spectroscopic characteristics. For example, the Field stains were mostly composed of Methylene blue, Azure A, and Brilliant cresyl blue. The Giemsa stains contained Azure C, Neutral red, Basic fuchsine, Pyronine derivatives, Brilliant cresyl blue, Indigo carmine, Alcian blue, Safranine O, Aniline blue and a wide range of unknown dyes. The Leishman stains appeared to be most spurious in composition as RGE-relevant dyes were found to have been replaced by irrelevant dyes such as Aniline blue, Neutral red, Thionine, Cresyl violet, Safranine, Phloxine, Azure C, Alizarin red, Brilliant cresyl blue and a range of unknown dyes. These findings are in keeping with the results obtained by Marshall et al. in evaluation of some commercial stains in the United Kingdom (Marshall et al., 1975a). Although levels of Azure B and Eosin Y were detected in some of the stains during HPLC assay despite the presence of spurious dyes, it became apparent that their concentrations were inadequate to provide an adequate ratio of Azure B to Eosin Y to produce the Romanowsky-Giemsa effect needed for satisfactory cytoplasmic and nuclear differentiation of malaria parasites. In this current study, excellent staining of parasites was obtained with the reference stain (Marshall et al., 1978), which yielded 25.0% and 32.8% of Azure B and Eosin Y respectively with an attendant Azure B to Eosin Y ratio of 0.8:1.0 A similarly performing stain was a Giemsa sample from the ANDi Laboratory, Lagos, which gave a ratio of 1.1. These findings are at variance with previous studies that demonstrated satisfactory staining with Azure B to Eosin Y ratio ranging from 5:1 – 10:1 (Wittekind, 1979). We, however, noted in this current study, a wide range of Azure B to Eosin Y ratio among the stain samples obtained from various laboratories and suppliers. It has been documented that commercial manufacturers of Romanowsky-Giemsa stains prefer an Azure B to Eosin Y ratio of 2:1 when formulating their stains (Marshall et al., 1978; Wittekind, 1979). In this current study, a number of stain samples exhibited Azure B to Eosin Y ratio around this commercially preferred ratio and higher. Furthermore, this study revealed that higher ratios of Azure B to Eosin Y does not necessarily confer excellent staining quality on the stains. This ratio was found to be quite high amongst the Field stains (yet none exhibited excellent staining), lower in the Giemsa stains and almost completely missing in the Leishman stains. These findings are a pointer to the fact that stain performance does not depend on Azure B to Eosin Y ratio alone. The Leishman stains performed most poorly. Previous workers have demonstrated that the Leishman stain is highly capricious both in composition and performance due to its method of preparation (Horobin, 2011; Marshall, 1978a; Bentley, 1980). In this current study, the Leishman stains were found to contain dyes other than Azure B and Eosin Y in order of concentration. Previous studies have demonstrated that combinations of Methylene blue and Eosin Y and Azure A and Eosin Y are unable to produce the RGE, resulting in the inability of the microscopist to differentiate the cytoplasm and the chromatin dots of malaria parasites (Horobin and Walter, 1987b; Wittekind, 1983; Wittekind and Gehring, 1985b). The finding of the presence of Eosin B in some of the stains in this current study may not be considered problematic as previous studies have demonstrated that Eosin B could be substituted for Eosin Y in the formulation of the Romanowsky-Giemsa stains (Friedrich et al., 1990b; Marshall et al., 1978, 1975c; Wilson, 1907; Wittekind, 1983).

Generally, the Rowmanowsky-Giemsa stains are procured by laboratories either as commercial ready-to-use solutions or as powders for reconstitution in appropriate solvents. In this study, we attempted to compare the dye compositions of commercial solutions versus those constituted from powder in-house against the reference stain of Marshall et al.(Marshall et al., 1975c). We have demonstrated that irrespective of mode of preparation, stains that have high amounts of Azure A, low Azure B, low Eosin Y and high Methylene blue combinations fail to give excellent staining results. This finding agrees with the results of previous Romanowsky-Giemsa stains researchers (Proctor and Horobin, 1985; Horobin and Walter, 1987a; Lubrano et al., 1977; Marshall, 1978a; Marshall et al., 1975c; Marshall and Galbraith, 1984). In addition, the higher the proportion of RGE-relevant dyes in the stains, the better the performance; stains that had pH lower than 7.0 and those with high sulphated ash percent tend to perform poorly. Specifically, among the Giemsa stains, we noted variation in the ratio of glycerol to methanol, the solvent used to constitute the stain. The higher the glycerol to methanol ratio, the poorer the performance of the stains. These findings tend to be in line with factors that have been previously demonstrated to influence performance of the Romanowsky-Giemsa stains (Marshall et al., 1975a, 1975c; Marshall, 1975a; Wittekind, 2008; Wittekind and Gehring, 1985). We have shown that minimum amounts of the RGE-relevant dyes are required to achieve the Romanowsky-Giemsa effect, which enables adequate cytoplasmic and nuclear colour differentiation in a stained malaria parasite. A minimum of 25% RGE-relevant dyes (Azure B and Eosin Y) was required for satisfactory staining. However, performance was pH-dependent and also affected by mode of stain preparation. For optimum performance, pH of 7.1-7.4 was required for the working solution, prepared from powder in-house. Although the proportion of Eosin Y present in the stains seems to be unimportant as found in this current study, the ratio of Azure B to Eosin Y is a crucial factor. We noted a ratio of at least 1.4:1.0 for commercially available stains to achieve satisfactory staining of malaria parasites. It appears that this ratio is also pH-dependent and could be easily achieved with in-house prepared stains. Previous studies have canvassed the need for laboratories to produce their stains locally (“in-house”) from scratch using commercial pure dyes (Marshall et al., 1975c, 1975a; Bentley, 1980). It must be noted that in this current study, the reference stain used was a Romanowsky-Giemsa stain formulated by Marshall et al (Marshall et al., 1975c) using commercial pure dyes. The stain was found to yield adequate amounts of Azure B and Eosin Y and the presence RGE-irrelevant dyes was minimal compared to the commercial stains sampled from laboratories. As demonstrated in this study, the dissolution of the RGE-relevant dyes in glycerol-methanolic solvent yields better stains, hence the use of Field and Leishman stains are not the best options for malaria microscopy. We recommend training laboratory personnel on the preparation of malaria microscopy stains from scratch using the pure RGE-relevant dyes and the evaluation of the stains for their composition. Perhaps the stain of Marshall et al. which was used as the reference Romanowsky-type stain in this study could be a possible choice as it can be readily prepared from individual dye components (Azure A, Azure B, Methylene Blue and Eosin Y). The malaria microscopist only needs to procure the individual powder dyes and dissolve in appropriate mixture of glycerol and methanol solvent and allow to brew. This would guarantee a stain of constant composition as previously demonstrated by the authors. There is urgent need to strengthen regulation of stains used for malaria microscopy and other IVDs both in Plateau State and in Nigeria at large. Further studies should be done to establish the role of impurities in the quality of colour imparted by the Romanowsky-type stains on malaria parasite and blood cells as we were unable to obtain enough powder stain samples from the facilities that participated in this study. The main independent predictors of staining quality seen in this study were the proportion of Azure B present in the stain; Azure B to Eosin Y ratio; and the solvent used to constitute the stain. Stains containing an Azure B proportion of 20% or more were over ten times more likely to produce satisfactory staining outcome; those with Azure B to Eosin Y ratio of 1.5 or more were about five times more likely to yield satisfactory staining; and stains constituted with alcoholic solvents were over twenty times more likely to produce satisfactory staining outcome. We noted that factors such as presence of RGE-antagonistic dyes, Eosin Y or B being the most abundant dye, presence of seven or more RGE-irrelevant dyes, and higher sulphated ash content were inimical to satisfactory staining outcome. In conclusion, we have, for the first time, profiled the physico-chemical characteristics of Romanowsky-type stains in Nigeria and elucidated on the factors that influence their performance in malaria parasite staining. The continuous stocking, supply and utilization of Romanowsky-type stains of variable composition, loaded with RGE-irrelevant dyes for malaria parasite cytoplasmic and nuclear differentiation (the so-called Romanowsky-Giemsa effect) in stained blood films has grievous consequences for malaria testing in Nigeria. As the NMEP presses towards malaria elimination in Nigeria, there is urgent need to develop and implement policies targeted at improving the quality of malaria microscopy reagents in both public and private laboratories to curtail either over- or under-estimation of the burden of malaria. This could possibly be achieved in collaboration with relevant regulatory agencies. The training of laboratory personnel on the preparation of a simple and stable Romanowsky-type stain from component dyes is pivotal for as long as microscopy remains the gold standard for malaria testing in this setting.

## Methods

### Study area

Plateau State is in North-central, Nigeria. The state has an estimated population of 4.5million people (Ria R. Ghai et al., 2016). Malaria is endemic in the state with a current prevalence among children aged 6 – 59 months estimated to be 13 – 22% by microscopy (Ilah Garba et al., 2016). This study was carried out in the 17 Local Government Areas in Plateau State. The LGAs are grouped into three senatorial districts-Plateau North (six LGAs), Plateau Central (five LGAs) and Plateau South (six LGAs). Plateau north consists mostly of metropolitan LGAs while the central and southern senatorial districts consist of majorly rural LGAs. Both public and private health facilities (including stand-alone laboratories) exist in all three senatorial districts of the State. There are 1492 health facilities in Plateau State. Among these are 1, 250, 220 and 22 primary, secondary and tertiary level facilities respectively. Overall, 237 (15.9%) facilities render malaria microscopy services in both public and private facilities across the three senatorial districts in Plateau State. The majority of primary health facilities use malaria RDT kits for testing.

### Study design and population

This was a facility-based onsite cross-sectional survey conducted among laboratories in public and private health facilities in Plateau State. It involved the collection of stain samples used for malaria microscopy (Field stain, Giemsa stain and Leishman stain). Laboratory services is provided by both public and private facilities in Plateau State, including malaria-testing services. Malaria microscopy is done mostly by secondary and tertiary level facilities. Most primary health facilities use malaria RDT kits for testing. There is no centralized stain distribution system, so each facility source for its own stain from suppliers in the open market. While most public facility laboratories use the WHO-recommended Giemsa stain, those in private facilities use other variants of Romanoski-type stains such as Field stain and Leishman stain for malaria microscopy. Moreover, while most public laboratories prefer to prepare their stains from commercial dye powder, majority of private laboratories prefer commercially prepared, ready-to-use stain solutions.

### Sample Size and Sampling Procedure

The sample size for laboratory facilities was calculated using the formula by Yamane (1967) (Yamane, 1967), which gave a minimum sample size of 144 laboratory facilities. A multi-stage sampling technique was used for sampling laboratory facilities offering malaria microscopy services across the three senatorial districts in Plateau State. A sampling frame was generated from data of laboratory facilities obtained from the Medical Laboratory Science Council of Nigeria (MLSCN), Plateau State Zonal office and Plateau State Ministry of Health. Only facilities that had a functional microscope and Romanowsky-type stains in current use were included in the study. Facilities that employed both malaria RDT kits and microscopy were excluded. Sampling was done by first selecting facilities on the basis of senatorial district ( Plateau North = 117, Plateau Central = 70, Plateau South = 50). Facilities were then further selected based on ownership either as “public” (n= 81) or “private” (n= 156). In every senatorial district, all public teaching hospitals and general hospitals were selected (census) while primary health centers and private facilities (including stand-alone laboratories) were selected by simple random sampling (balloting) until the minimum sample size was completed. In all, 92 facilities had an adequate quantity of stains to offer for the study.

### Data Collection

An interviewer-administered stain information questionnaire was used to capture relevant data, including date, stain type, batch number, expiry date, mode of stain preparation, form of stain and name of stain manufacturer. Twenty milliliters (20mL) of the stain currently used for malaria microscopy was collected in a labeled sterile universal bottle alongside 20mL of the water used for dilution (for those using Giemsa stain). The stain solution was immediately kept in a locked mobile cupboard to prevent stain exposure to light. For facilities, that prepared their own stains solution from powder, 20grams of the powder form was also collected. All stain samples and diluents were carefully labelled with a facility number.

### Laboratory Techniques

The dye components of the stains were determined according to the methods of the Biological Stains Commission (Penney et al., 2002), using high performance liquid chromatography (HPLC) and ultra violet spectrophotometry. The Agilent Series 1050 HPLC system with Agilent Chemstation software was used for HPLC assay, Agilent 8453 UV-VIS spectrophotometer was used for spectrophotometric analysis and Agilent 7697A gas chromatography analyzer was used for stains solvent analysis. The method of stain performance evaluation described by (Marshall et al., 1975a)) was used: Venous blood was obtained in 5ml EDTA container from three subjects-hematological normal, megaloblastic anemia and acute granulocytic leukemia. Blood sample confirmed to be positive for *Plasmodium falciparum* in 5mL EDTA container was obtained and used to make thick and thin blood films on a clean, grease-free glass slide and air-dried according to the standard method (WHO, 2016b). Methanol fixing was done for films meant for Giemsa and Field stains staining. The number of slides made for each sample corresponded to the number of stain samples collected and the reference stain. The dried films were stained with the working solutions of the sampled stains according to standard hematology procedure, dried and examined microscopically by WHO-certified expert microscopists using a pre-calibrated microscope.

### Data processing and analysis

The main outcome variable was quality of colour imparted on malaria parasites in the blood smear, which was defined as “satisfactory” if the colour was graded as “excellent”, “fair” or “good” and “unsatisfactory” if the colour was graded as “poor” by the WHO-certified microscopists. The main independent variables were stain type, the mode of stain preparation, proportion of methylene blue, proportion of Azure B, proportion of Eosin Y, Azure B:Eosin Y ratio, stain pH, proportion of RGE-relevant dye, and type of solvent used. Data was entered using Microsoft Word Excel version 2016 and analyzed using R statistical software. We summarized categorical variables using frequencies and percentages, while numerical variables were summarized using the median and interquartile range. Comparison of percentages between outcome and explanatory variables was done using Pearson’s Chi-square statistic and Fisher’s exact test for categorical variables and Kruskal-Wallis rank sum test for continuous variables with p-values <0.05 considered to be statistically significant. To estimate the strength of the association between explanatory variables and the outcome variable, we calculated prevalence odds ratios with their corresponding 95% confidence intervals and p-values. We used a simple logistic regression model to demonstrate the effect of individual determinants on the likelihood of good quality staining. By estimating the crude prevalence odds ratios and their associated 95% confidence intervals, we showed the strength and direction of the association between each predictor and the likelihood of “satisfactory” staining outcome. We used a directed acyclic graph (DAG) (Textor et al., 2016) to illustrate the relationship between the outcome variable and the independent variables (**Supplementary Fig 1**) and a stepwise multivariable binary logistic regression approach (Akar et al., 2025; Tuboi et al., 2007) to assess the effects of multiple predictors of satisfactory staining quality simultaneously. Here, we used an Akaike Information Criterion-based stepwise model to systematically eliminate the predictors. We corrected the output of the final model for highly colinear predictors by removing variables with high variance inflation factor (VIF > 5). We determined the independence of each predictor using the likelihood ratio test. We fitted a final binary logistic regression model (estimated using maximum likelihood) to predict “satisfactory” staining outcome with proportion of Azure B >20%, proportion of Methylene Blue >25%, Eosin B or Y as most prominent dye, Azure B to Eosin Y ratio <2, presence of antagonistic dyes, number of unknown dyes <7, number of RGE-irrelevant dyes >7, pH, solvent type, and sulphated ash percent category. The model’s explanatory power was moderate (Tjur’s R2 = 0.298).’ Standardized parameters were obtained by fitting the model on standardized dataset version. 95% Confidence Intervals (CIs) and p-values were computed using a Wald z-distribution approximation. This simultaneous modeling allows the calculation of adjusted prevalence odds ratios (aOR) as a measure of the contribution of individual predictors after adjusting for other factors included in the model.

## Supporting information

Akar et al Supplementary files

## Data Availability

All data produced in the present study are available upon reasonable request to the authors.

## Acknowledgement

We wish to appreciate Dr John Kernan and Prof Richard Horobin, both of the Biological Stains Commission for providing useful literature for this work. We appreciate the team of laboratory technologists at the Department of Chemistry, Faculty of Science, Bowen University, Iwo, Osun State, Nigeria for their insightful guidance in developing the laboratory analysis protocols used in this study.

## Contributors

S.E.A., M.D., A.K., I..O., O.O., and W.O. contributed with literature review, study design and development of study tool. M.D. and A.K. supervised the field activities. S.E, M.D., A.K., O.A., O.O. and W.O. did data analysis and result interpretation. O.A, G.N. and W.T. provided relevant operational and field information and guidelines to enable implementation of the study. All authors were involved in manuscript review for publication

## Ethical approval

Ethical approval for the study was obtained from the University of Ibadan/University College Hospital Ethics Committee (Ref:UI/EC/17/0352), Plateau State Ethical Review Committee (Ref: MOH/MIS/202/VOL.T/X), Plateau State Hospitals Management Board (ADM/554/vol.1/141) and Jos University Teaching Hospital Ethics Committee (Ref: JUTH/DCS/ADM/127/XXVII/704).

## Provenance and peer review

Not commissioned; externally peer reviewed.

## Data availability statement

The dataset used for this analysis is available on request.

## Open access

This is an open access article distributed in accordance with the Creative Commons Attribution 4.0 Unported (CC BY 4.0) license, which permits others to copy, redistribute, remix, transform and build upon this work for any purpose, provided the original work is properly cited, a link to the licence is given, and indication of whether changes were made. See: https://creativecommons.org/licenses/by/4.0/.

## Supplementary Information

### Supplementary Figures

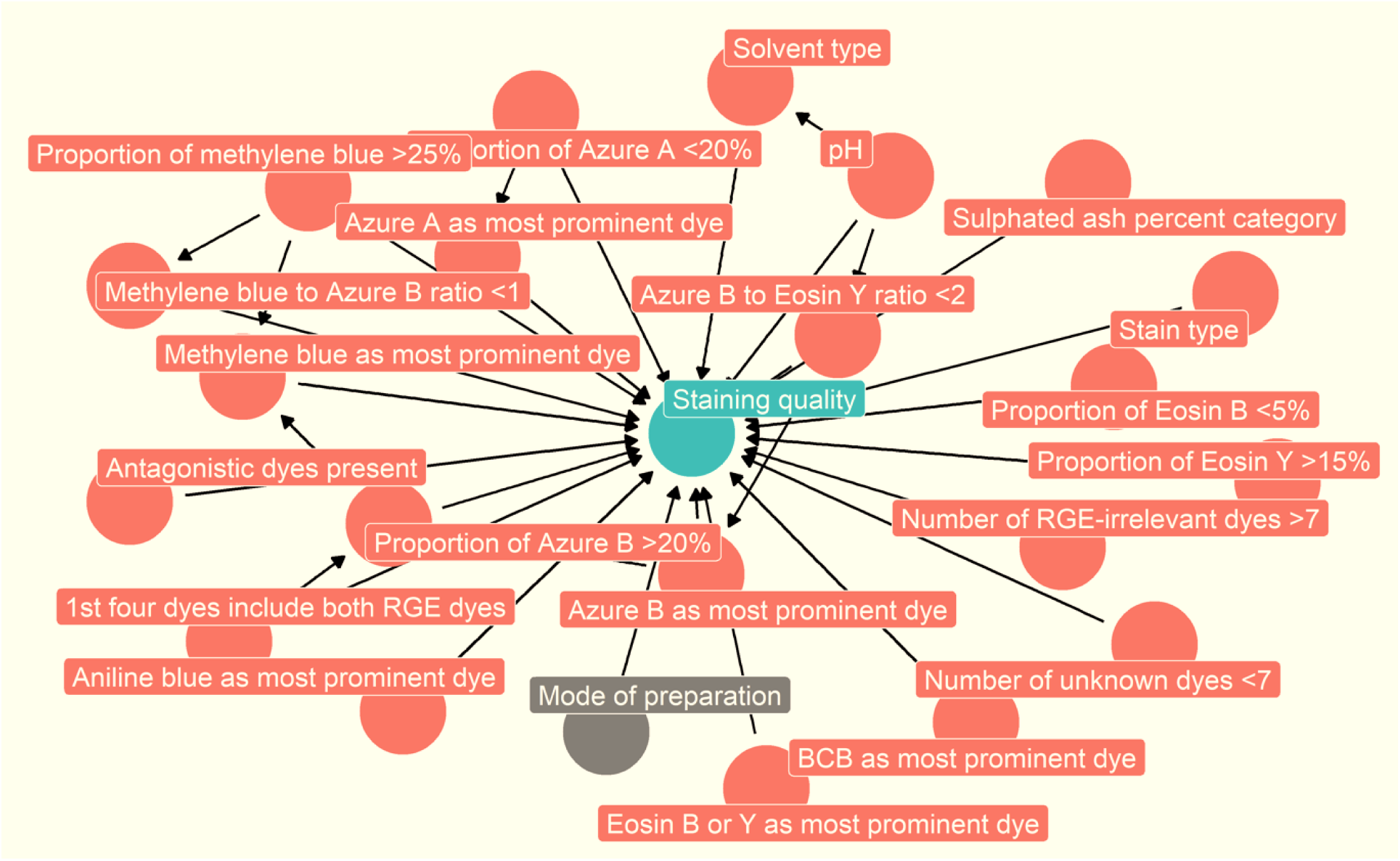

**Supplementary Fig 2.**
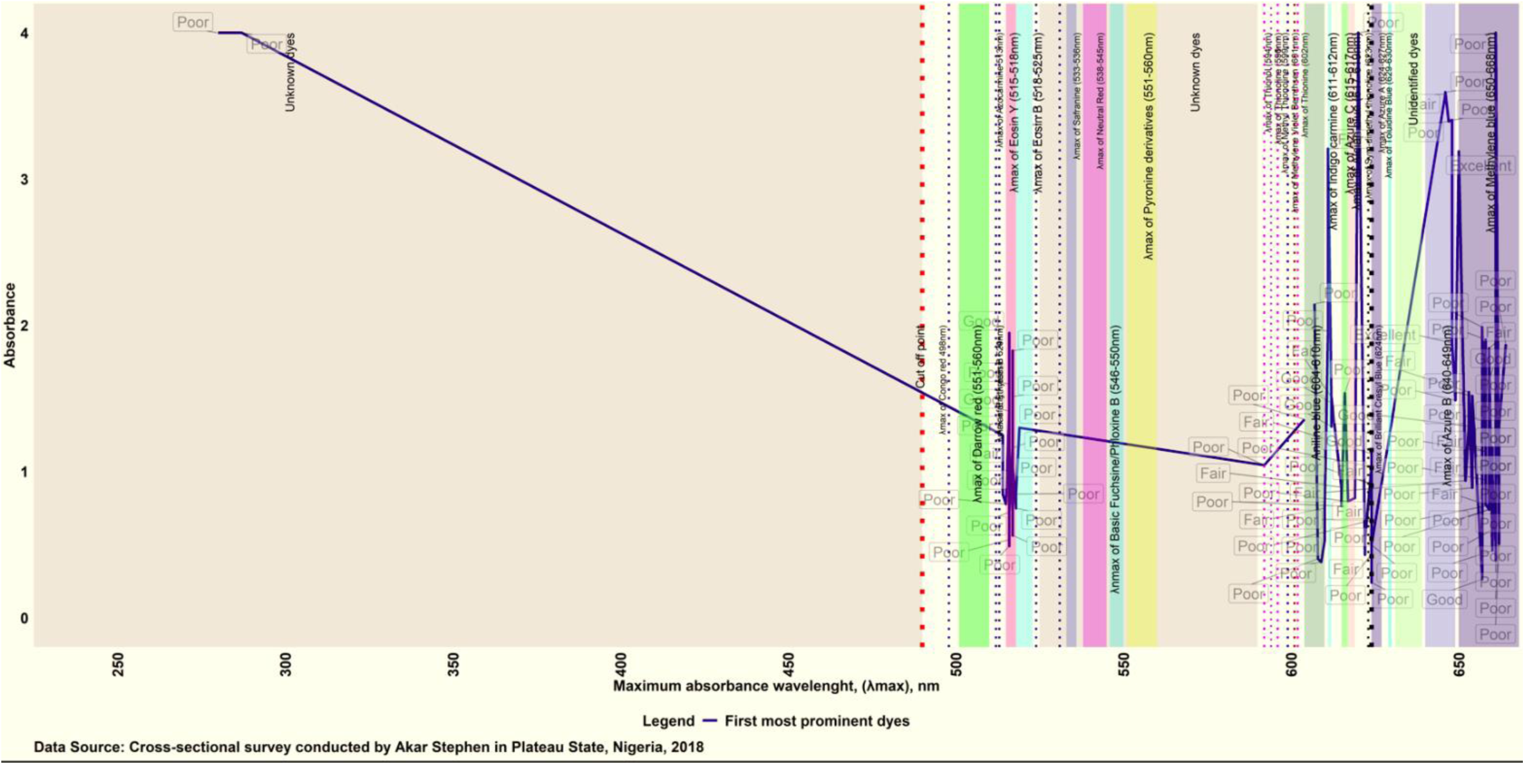
Spectroscopic characterisitcs of the first prominent dye components of Romanowski-type stains used for malaria microscopy in Plateau state, Nigeria disaggregated by staining colour grade

**Supplementary Fig 3.**
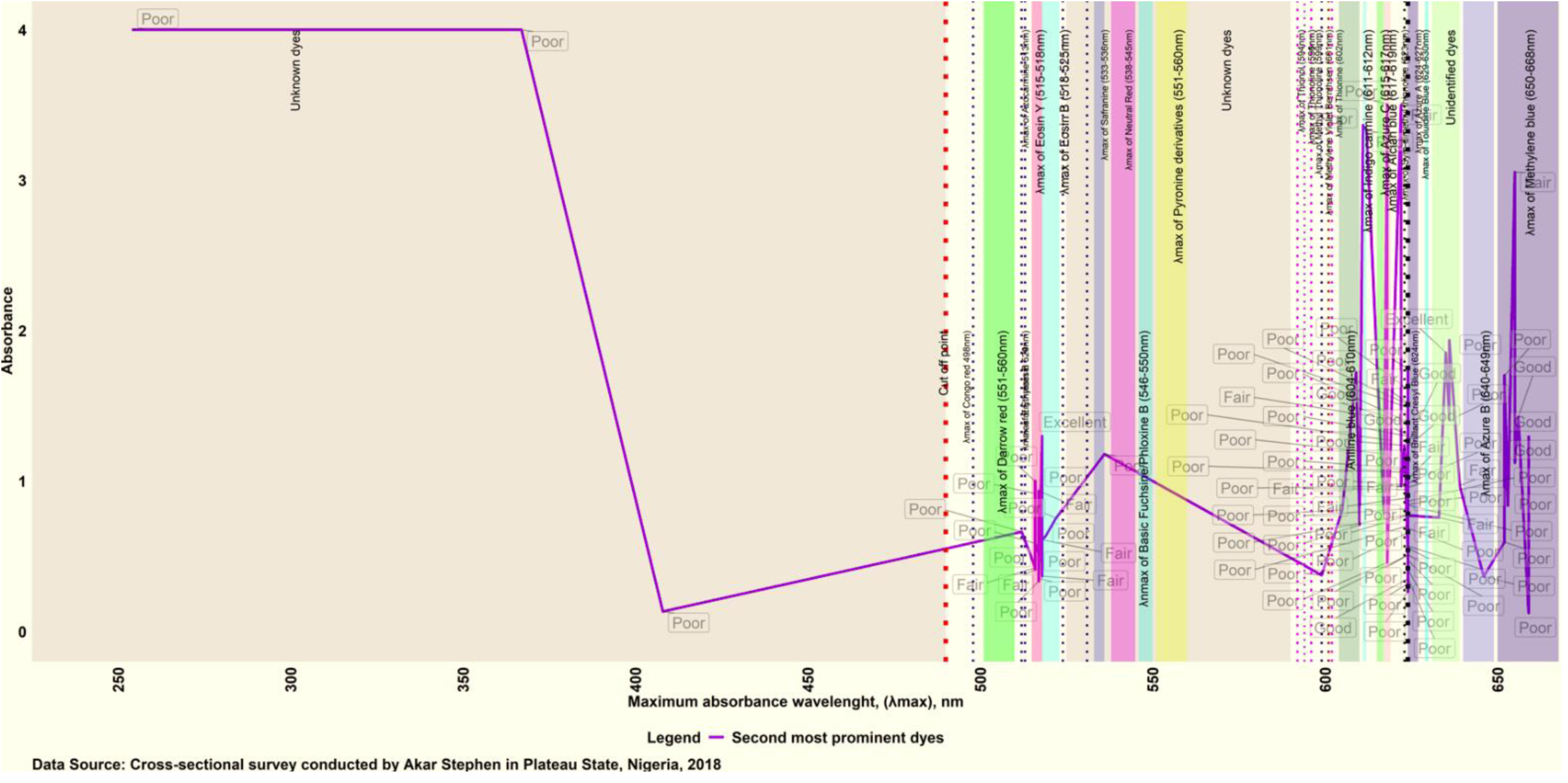
Spectroscopic characteristics of second most prominent dye components of Romanowski-type stains used for malaria microscopy in Plateau state, Nigeria disaggregated by staining colour grade

**Supplementary Fig 4.**
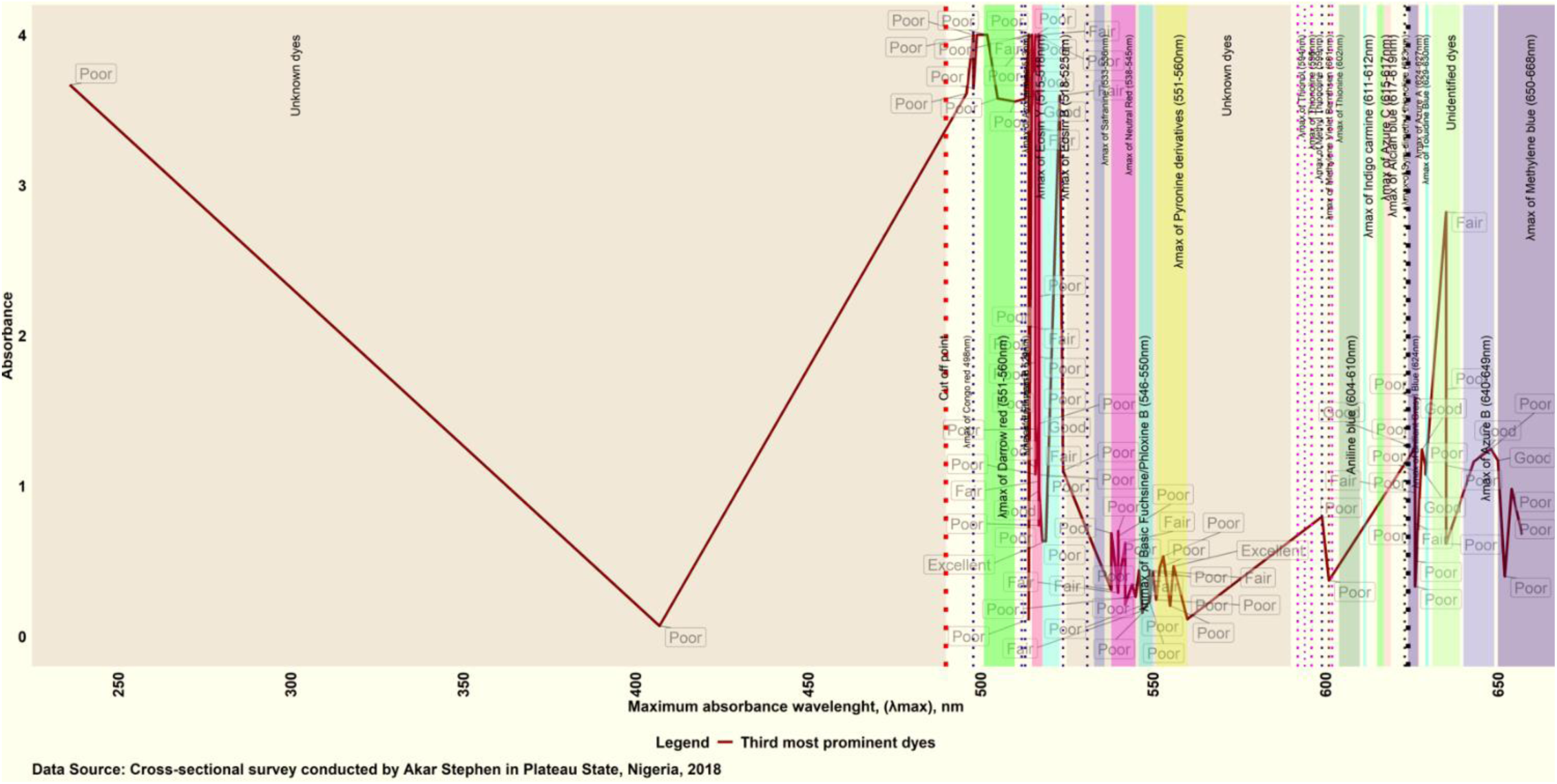
Spectroscopic characteristics of third most prominent dye components of Romanowski-type stains used for malaria microscopy in Plateau state, Nigeria disaggregated by staining colour.

**Supplementary Fig 5.**
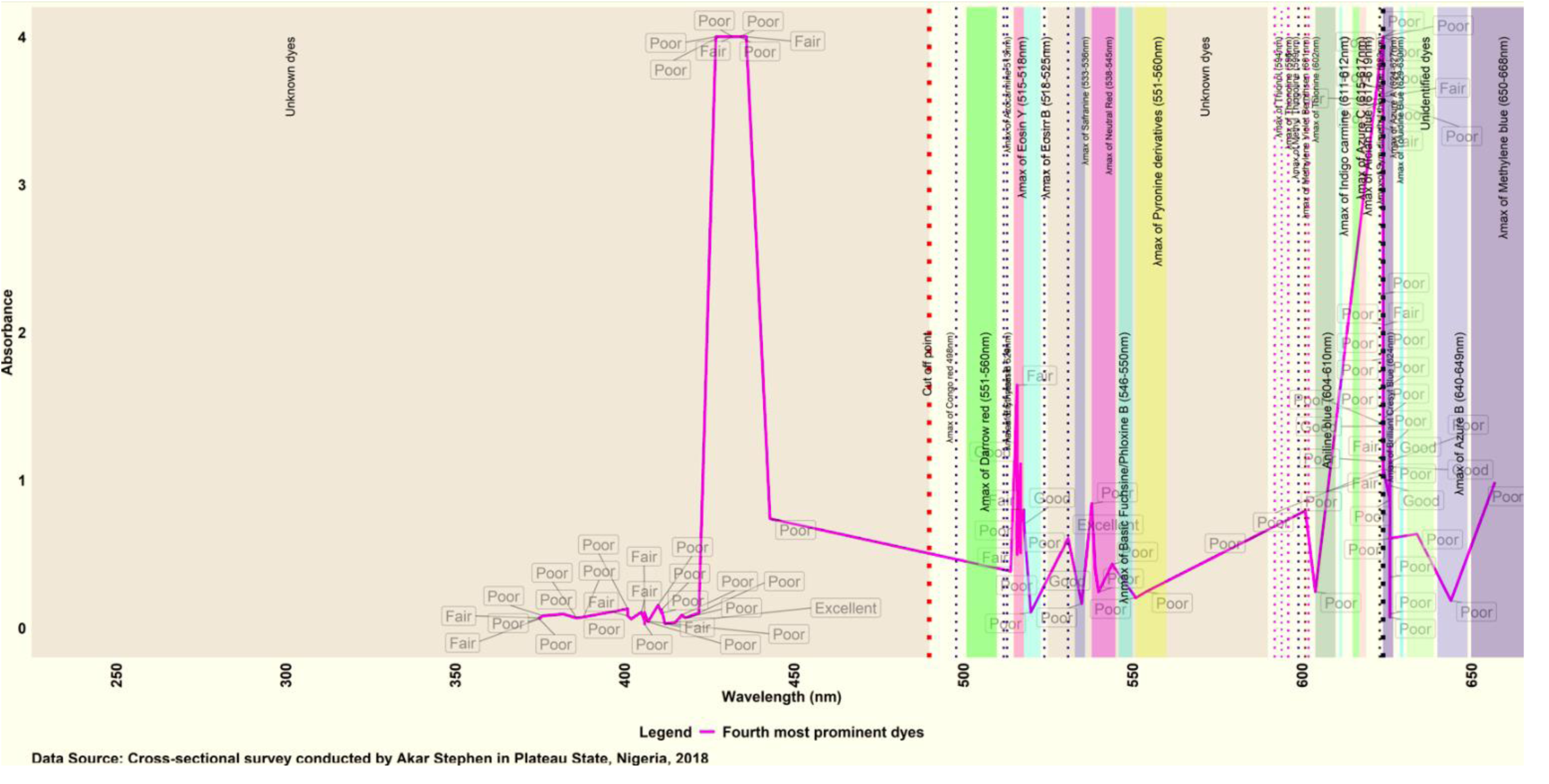
Spectroscopic characterisitcs of fourth most prominent dye components of Romanowski-type stains used for malaria microscopy in Plateau state, Nigeria disaggregated by staining colour grade

### Supplementary Tables

**Table 1.** Identification of dye species present in Field stains solution used for malaria microscopy in Plateau State Nigeria based on the maximum absorbance wavelengths of specific dye species determined by ultra-violet spectrophotometric assay.

| Stain ID | Batch number | Expiry date | Identity of the four most prominent dyes based on value of maximum absorbance wavelengths (λmax) of the specific dye components |  |  |  |
| --- | --- | --- | --- | --- | --- | --- |
| Marshall and Lewis stain (reference) |  |  |  |  |  |  |
| ML | Yes | Yes | Azure B (650nm) | Eosin B/Eosin Y (518) | Pyronine derivative (556nm) | Unknown dye (419) |
| BEMA Scientific: 11 (28.9%) |  |  | λmax = 652 – 662nm | λmax = 624nm | λmax = 626nm | λmax = 498-499nm, 516 - 517nm |
| Field 1 | No | No | Methylene blue | Brilliant cresyl blue | Azure A | Eosin B/Eosin Y |
| Field 7 | No | No | Methylene blue | Brilliant cresyl blue | Azure A | Congo red (498nm) |
| Field 15 | No | No | Methylene blue | Brilliant cresyl blue | Azure A | Eosin B/Eosin Y |
| Field 19 | No | No | Methylene blue | Brilliant cresyl blue | Azure A | Congo red (499nm) |
| Field 48 | No | No | Methylene blue | Brilliant cresyl blue | Azure A | Eosin B/Eosin Y |
| Field 53 | No | No | Methylene blue | Brilliant cresyl blue | Azure A | Eosin B/Eosin Y |
| Field 72 | No | No | Methylene blue | Brilliant cresyl blue | Azure A | Eosin B/Eosin Y |
| Field 80 | No | No | Methylene blue | Brilliant cresyl blue | Azure A | Azocarmine G (510nm) |
| Field 84 | No | No | Methylene blue | Brilliant cresyl blue | Azure A | Eosin B/Eosin Y |
| Field 85 | No | No | Methylene blue | Brilliant cresyl blue | Azure A | Eosin B/Eosin Y |
| Field 116 | No | No | Methylene blue | Brilliant cresyl blue | Azure A | Eosin B/Eosin Y |
| Ranjo Medix Product: 9 (23.7%) |  |  | λmax = 651 – 661nm | λmax = 624, 636nm | λmax = 626nm | λmax = 502nm, 515 – 517nm |
| Field 3 | No | No | Methylene blue | Brilliant cresyl blue | Azure A | Eosin B/Eosin Y |
| Field 16 | No | No | Methylene blue | Brilliant cresyl blue | Azure A | Darrow/Congo red (502nm) |
| Field 21 | No | No | Methylene blue | Brilliant cresyl blue | Azure A | Eosin B/Eosin Y |
| Field 30 | No | No | Methylene blue | Brilliant cresyl blue | Azure A | Eosin B/Eosin Y |
| Field 31 | No | No | Methylene blue | Brilliant cresyl blue | Azure A | Eosin B/Eosin Y |
| Field 40 | No | No | Methylene blue | Brilliant cresyl blue | Azure A | Eosin B/Eosin Y |
| Field 125 | No | No | Methylene blue | Brilliant cresyl blue | Azure A | Eosin B/Eosin Y |
| Field 126 | No | No | Methylene blue | Unknown dye (636nm) | Azure A | Eosin B/Eosin Y |
| Field 145 | No | No | Methylene blue | Azure C derivative | Azure A | Eosin B/Eosin Y |
| AFIS Biochemicals: 4 (10.5%) |  |  | λmax = 646nm, 653–654nm | λmax = 618-622nm , 624nm | λmax = 626, 635nm | λmax = 496nm, 516–517nm, 523nm |
| Field 14 | No | No | Methylene blue | Azure C derivative | Azure A | Eosin Y/Eosin B |
| Field 71 | No | No | Methylene blue | Azure C derivative | Azure A | Eosin Y/Eosin B |
| Field 79 | No | No | Azure B (646nm) | Azure C | Azure A | Erythrosin B (523nm) |
| Field 97 | No | No | Azure B (646nm) | Brilliant Cresyl Blue | Unknown dye | Congo red (496nm) |
| Trust Chemical Laboratories 3 (7.9%) |  |  | λmax = 658 - 661 | λmax = 624nm, 636nm | λmax = 518nm, 624, 626nm | λmax = 515 – 517nm |
| Field 0 | Yes | Yes | Methylene blue | Azure C derivative | Eosin B/Eosin Y | Eosin B/Eosin Y |
| Field 28 | Yes | Yes | Methylene blue | Azure C derivative | Brilliant cresyl blue | Eosin B/Eosin Y |
| Field 88 | Yes | Yes | Methylene blue | Unknown dye | Azure A | Eosin B/Eosin Y |
| Field 152 | Yes | Yes | Methylene blue | Azure A | Azure C derivative | Eosin B/Eosin Y |
| **Suzinal Chemicals: 2 (5.3%)** |  |  | **$\lambda_{\max} = 658\text{nm}$** | **$\lambda_{\max} = 624\text{nm}$** | **$\lambda_{\max} = 626\text{nm}$** | **$\lambda_{\max} = 517 - 518\text{nm}$** |
| Field 33 | No | No | Methylene blue | Brilliant cresyl blue | Brilliant cresyl blue | Eosin Y/Eosin B |
| Field 69 | No | No | Methylene blue | Brilliant cresyl blue | Brilliant cresyl blue | Eosin Y/Eosin B |
| **Chemlight: 1 (2.6%)** |  |  |  |  |  |  |
| Field 57 | Yes | Yes | Azure B (649nm) | Brilliant cresyl blue | Brilliant cresyl blue (626nm) | Eosin Y/Eosin B (517nm) |
| **Ferntec: 1 (2.6%)** |  |  |  |  |  |  |
| Field 64 | Yes | Yes | Methylene blue (660nm) | Brilliant cresyl blue | Brilliant cresyl blue (626nm) | Darrow red (506nm) |
| **God's Defence Medical Laboratory: 1 (2.6%)** |  |  |  |  |  |  |
| Field 50 | No | No | Azure B (648nm) | Aniline blue (611nm) | Toluidine blue (629nm) | Azocarmine G (513nm) |
| **Kaycee Chemicals: 1 (2.6%)** |  |  |  |  |  |  |
| Field 23 | No | No | Azure B (647nm) | Indigo carmine (612nm) | Brilliant cresyl blue (628nm) | Eosin Y derivative (514nm) |
| **Kenac Reagents: 1 (2.6%)** |  |  |  |  |  |  |
| Field 6 | No | No | Methylene blue (660nm) | Brilliant cresyl blue (624nm) | Brilliant cresyl blue (626nm) | Eosin Y/Eosin B (516nm) |
| **Kenol Pharmacy and Laboratories: 1 (2.6%)** |  |  |  |  |  |  |
| Field 59 | Yes | Yes | Methylene blue (653nm) | Brilliant cresyl blue (624nm) | Brilliant cresyl blue (626nm) | Eosin Y (515nm) |
| **Lab Tech Chemicals: 1 (2.6%)** |  |  |  |  |  |  |
| Field 54 | Yes | Yes | Indigo carmine/aniline blue (611nm) | Unknown dye (408nm) | Unknown dye (290nm) | Congo red (498nm) |
| **P.Park Scientific** |  |  |  |  |  |  |
| Field 138 | Yes | Yes | Alcian blue (617nm) | Unknown dye (367nm) | Unknown dye (473nm) | Eosin B/Eosin Y (517nm) |

**Table 2.** Identification of dye species present in Giemsa stains solution used for malaria microscopy in Plateau State Nigeria based on the maximum absorbance wavelengths of specific dye species determined by ultra-violet spectrophotometric assay.

| Stain ID | Batch number | Expiry date | Identity of the four most prominent dyes based on value of maximum absorbance wavelengths ($\lambda_{\max}$) of the specific dye components present in the aqueous stain solution | | | |
| --- | --- | --- | --- | --- | --- | --- |
| | | | $\lambda_{\max}$ = 615nm, 624nm, 514 – 517nm | $\lambda_{\max}$ = 516nm, 609nm, 622, 624nm | $\lambda_{\max}$ = 407nm, 540-549nm, 550-553nm, 654nm | $\lambda_{\max}$ = 401-412nm, 626nm |
| **Marshall and Lewis stain (reference)** |  |  |  |  |  |  |
| ML | Yes | Yes | Azure B (650nm) | Eosin B/Eosin Y (518) | Pyronine derivative (556nm) | Unknown dye (419) |
| **BEMA Scientific: 6 (16.7%)** |  |  |  |  |  |  |
| Giemsa4 | No | No | Azure C (615nm) | Eosin B/Eosin Y (516) | Neutral red (540nm) | Unknown dye (412nm) |
| Giemsa 20 | No | No | Brilliant cresyl blue(624nm) | Eosin B/Eosin Y (516) | Methylene blue (654nm) | Azure A(626nm) |
| Giemsa22 | No | No | Eosin B/Eosin Y (517nm) | Brilliant cresyl blue(624nm) | Basic fuchsine (550nm) | Unknown dye (407nm) |
| Giemsa48 | No | No | Eosin Y (514nm) | Sym-dimethyl thionine (622nm) | Pyronine derivative (553nm) | Unknown dye (401nm) |
| Giemsa97 | No | No | Unknown dye (280nm) | Aniline blue (609nm) | Unknown dye (407nm) | Nil |
| Giemsa107 | No | No | Brilliant cresyl blue (624nm) | Eosin B/Eosin Y (516nm) | Phloxine B (549nm) | Unknown dye (406nm) |
| **Trust Chemical Laboratories: 5 (13.9%)** | | | $\lambda_{\max}$ = 517nm, 612nm, 624nm | $\lambda_{\max}$ = 517nm, 624nm, 652-655nm | $\lambda_{\max}$ = 517nm, 542nm, 626nm, 629nm, 648nm | $\lambda_{\max}$ = 406nm, 520nm, 626nm |
| Giemsa99 | Yes | Yes | Eosin B/Eosin Y (517nm) | Brilliant cresyl blue (624) | Azure B (648nm) | Azure A (626nm) |
| Giemsa104 | Yes | Yes | Brilliant cresyl blue (624nm) | Methylene blue (652nm) | Azure A (626nm) | Eosin B (520nm) |
| Giemsa118 | Yes | Yes | Eosin B/Eosin Y (517nm) | Brilliant cresyl blue (624nm) | Toluidine blue (629nm) | Azure A (626nm) |
| Giemsa149 | Yes | Yes | Indigo carmine (612nm) | Eosin B/Eosin Y (517nm) | Neutral red (542nm) | Unknown dye (406nm) |
| Giemsa150 | Yes | Yes | Brilliant cresyl blue (624nm) | Methylene blue (655nm) | Eosin B/Eosin Y (517nm) | Azure A(626nm) |
| **Kem Light Reagents: 3 (8.3%)** | | | $\lambda_{\max}$ = 516, 607nm, 620nm | $\lambda_{\max}$ = 536, 624nm, 652nm | $\lambda_{\max}$ = 524nm, 635nm, 643nm | $\lambda_{\max}$ = 411nm, 538nm , 626nm |
| Giemsa76 | Yes | Yes | Eosin B/Eosin Y (516nm) | Brilliant cresyl blue (624nm) | Erythrosin B (524nm) | Azure A (626nm) |
| Giemsa133 | Yes | Yes | Aniline blue (607nm) | Methylene blue (652nm) | Unknown dye (635nm) | Unknown dye (411nm) |
| Giemsa 139 | Yes | Yes | Alcian blue (620nm) | Safranin O (536nm) | Azure B derivative (643nm) | Neutral red (538nm) |
| **ANDi Quality Assurance Laboratory, Lagos: 2 (5.6%)** | | | $\lambda_{\max}$ = 519nm, 648nm | $\lambda_{\max}$ = 623nm, 635nm | $\lambda_{\max}$ = 519nm, 635nm | |
| Giemsa0 | Yes | Yes | Azure B (648nm) | Safranin O (635nm) | Eosin B (519nm) | Ethyl eosin (531nm) |
| Giemsa132 | Yes | Yes | Eosin B (519nm) | Sym-dimethyl thionine (623nm) | Unknwon (635nm) | Methylene blue (657nm) |
| **Apex Laboratories Reagents: 2 (5.6%)** | | | $\lambda_{\text{max}} = 516\text{nm}, 610\text{nm}$ | $\lambda_{\text{max}} = 517\text{nm}, 624\text{nm}$ | $\lambda_{\text{max}} = 545\text{nm}, 650\text{nm}$ | $\lambda_{\text{max}} = 388\text{nm}, 626\text{nm}$ |
| Giemsa55 | No | No | Eosin B/Eosin Y (516nm) | Brilliant cresyl blue (624nm) | Azure B (650nm) | Azure A (626nm) |
| Giemsa141 | No | No | Aniline blue (610nm) | Eosin B/Eosin Y (517nm) | Neutral red (545nm) | Unknown dye (388nm) |
| **Unknown Manufacturer: 2 (5.6%)** | | | $\lambda_{\text{max}} = 624\text{nm}$ | $\lambda_{\text{max}} = 653\text{—}659\text{nm}$ | $\lambda_{\text{max}} = 514, 624\text{nm}$ | $\lambda_{\text{max}} = 517, 626\text{nm}$ |
| Giemsa41 | No | No | Brilliant cresyl blue (624nm) | Methylene blue (659nm) | Eosin Y (514nm) | Azure A (626nm) |
| Giemsa157 | No | No | Brilliant cresyl blue (624nm) | Methylene blue (653nm) | Brilliant cresyl blue (624nm) | Eosin Y/Eosin B (517nm) |
| **Afis Biochemicals: 1 (2.8%)** |  |  |  |  |  |  |
| Giemsa 65 | No | No | Methylene blue (654nm) | Brilliant cresyl blue (624nm) | Azure A (626nm) | Neutral red (544nm) |
| **Antek Diagnostics Ltd, UK: 1 (2.8%)** |  |  |  |  |  |  |
| Giemsa 120 | Yes | Yes | Indigo carmine (612nm) | Methylene blue (659nm) | Toluidine blue (629nm) | Neutral red (539nm) |
| **Ferntec: 1 (2.8%)** |  |  |  |  |  |  |
| Giemsa 91 | Yes | Yes | Aniline blue (606nm) | Eosin B (519nm) | Safranin O (533nm) | Unknown dye (406nm) |
| **Hembarge Chemical Company: 1 (2.8%)** |  |  |  |  |  |  |
| Giemsa 154 | Yes | Yes | Methylene blue (664nm) | Alcian blue (619nm) | Azure A (626nm) | Unknown dye (418nm) |
| **Jos University Hospital Laboratory: 1 (2.8%)** |  |  |  |  |  |  |
| Giemsa 2 | Yes | Yes | Methylene blue (657nm) | Alcian blue (618nm) | Azure A (626nm) | Eosin Y/Eosin B (517nm) |
| **Kaycee Chemicals: 1 (2.8%)** |  |  |  |  |  |  |
| Giemsa 84 | No | No | Eosin Y/Eosin B (516nm) | Brilliant cresyl blue (624nm) | Pyronine B (555nm) | Pyronine derivative (551nm) |
| **Kermel: 1 (2.8%)** |  |  |  |  |  |  |
| Giemsa 121 | Yes | Yes | Methylene blue (662nm) | Alcacian blue (618nm) | Azure A (626nm) | Eosin Y/Eosin B (517nm) |
| **Lab Tech Chemicals: 1 (2.8%)** |  |  |  |  |  |  |
| Giemsa 61 | Yes | Yes | Indigo carmine (611nm) | Methylene blue (655nm) | Azure A (635nm) | Eosin B/Eosin Y (516nm) |
| **Medicom Laboratories: 1 (2.8%)** |  |  |  |  |  |  |
| Giemsa 8 | Yes | Yes | Eosin Y (515nm) | Brilliant cresyl blue (624nm) | Thionine dimer (565nm) | Azure B derivative (644nm) |
| **Merck: (2.8%)** |  |  |  |  |  |  |
| Giemsa 62 | Yes | Yes | Eosin Y (514nm) | Alcian blue (617nm) | Basic fuchsine (546nm) | Unknown dye (376nm) |
| **Plateau State Specialist Hospital Laborator: (2.8%)** |  |  |  |  |  |  |
| Giemsa 25 | Yes | Yes | Methylene blue (657nm) | Brilliant cresyl blue (624nm) | Azure A (626nm) | Eosin Y (514nm) |
| **Raymoond A Lamb: (2.8%)** |  |  |  |  |  |  |
| Giemsa 98 | Yes | Yes | Aniline blue (604nm) | Eosin B/Eosin Y (516nm) | Neutral red (540nm) | Unknown dye (402nm) |
| **S. Merck Chemicals: 1 (2.8%)** |  |  |  |  |  |  |
| Giemsa 119 | Yes | Yes | Indigo carmine (612nm) | Methylene blue (656nm) | Azure A (628nm) | Eosin B/Eosin Y (518nm) |

**Table 3.** Identification of dye species present in Leishman stains solution used for malaria microscopy in Plateau State Nigeria based on the maximum absorbance wavelengths of specific dye species determined by ultra-violet spectrophotometric assay.

| Stain ID | Batch number | Expiry date | Identity of the four most prominent dyes based on value of maximum absorbance wavelengths (λmax) of the specific dye components present in the aqueous stain solution |  |  |  |
| --- | --- | --- | --- | --- | --- | --- |
|  |  |  | λmax = 517nm, 608 – 609nm | λmax = 517nm, 599-605nm | λmax = 542nm, 599-601nm | λmax = 386nm, 542nm, 601nm |
| Marshall and Lewis stain (reference) |  |  |  |  |  |  |
| ML | Yes | Yes | Azure B (650nm) | Eosin B/Eosin Y (518) | Pyronine derivative (556nm) | Unknown dye (419) |
| BEMA Scientific: 3 (15.0%) |  |  |  |  |  |  |
| Leishman9 | No | No | Aniline blue/Indigo carmine (609nm) | Eosin B/Eosin Y (517) | Neutral red (542nm) | Unknown dye (386nm) |
| Leishman70 | No | No | Eosin B/Eosin Y (517nm) | Aniline blue (605nm) | Methyl thionine (599nm) | Methylene violet Bernthsen (601nm) |
| Leishman106 | No | No | Aniline blue (608 - 609nm) | Methyl thionoline (599nm) | Methylene violet Bernthsen (601nm) | Neutral red (542nm) |
| Kaycee Chemicals: 3 (15.0%) |  |  | λmax = 516nm, 610nm, 622nm | λmax = 516-518nm, 618nm | λmax = 538nm, 547nm, 551nm | λmax = 375-382nm |
| Leishman25 | No | No | Aniline blue (610nm) | Eosin B/Eosin Y (518nm) | Neutral red (538nm) | Unknown dye (375nm) |
| Leishman60 | No | No | Eosin B/Eosin Y (516nm) | Alcian blue (618nm) | Pyronine derivative (551nm) | Unknown dye (382nm) |
| Leishman63 | No | No | Sym-dimethyl thionine (622nm) | Eosin B/Eosin Y (516nm) | Unknown (547nm) | Unknown dye (375nm) |
| Medicom Laboratories: 2 (10.0%) |  |  | λmax = 516-517nm | λmax = 624nm | λmax = 549nm, 635nm | λmax = 405nm, 626nm |
| Leishman51 | Yes | Yes | Eosin B/Eosin Y (516nm) | Brilliant cresyl blue (624nm) | Basic fuchsine (549nm) | Unknown dye (405nm) |
| Leishman86 | Yes | Yes | Eosin B/Eosin Y (517nm) | Azure C derivative (624nm) | Unknown (635nm) | Azure A (626nm) |
| Sigma Aldreich, Germany: 2 (10.0%) |  |  | λmax = 518nm, 610nm | λmax = 624nm | λmax = 626nm, 657nm | λmax = 535 – 554nm |
| Leishman155 | Yes | No | Eosin B (518nm) | Brilliant cresyl blue (624nm) | Methylene blue (657nm) | Pyronine derivative (554nm) |
| Leishman98 | Yes | Yes | Brilliant cresyl blue (624nm) | Azure B (646nm) | Azure A (626nm) | Safranine (535nm) |
| Ferntec ASN: 1 (5.0%) |  |  |  |  |  |  |
|  | Yes | Yes | Brilliant cresyl blue(624nm) | Eosin Y/Eosin B (517nm) | Basic fuchsine (547nm) | Unknown dye (375nm) |
| Hembarge Chemicals Company: 1 (5.0%) |  |  |  |  |  |  |
| Leishman132 | Yes | Yes | Azure C (616nm) | Eosin Y/Eosin B (516nm) | Neutral red (538nm) | Unknown dye (411nm) |
| Javic: 1 (5.0%) |  |  |  |  |  |  |
| Leishman<br>26 | No | No | Eosin Y/Eosin B (516nm) | Aniline blue/Azure C<br>(610nm) | Pyronine<br>derivative(552nm) | Unknown dye (410nm) |
| **Kem Light Laboratories: 1 (5.0%)** |  |  |  |  |  |  |
| Leishman<br>89 | No | No | Alizarin red (592nm) | Azure C (616nm) | Neutral red (540nm) | Unknown dye (422nm) |
| **Lab Tech Chemicals: 1 (5.0%)** |  |  |  |  |  |  |
| Leishman<br>115 | Yes | Yes | Unknown dye (287nm) | Unknown dye (254nm) | Unknown dye (236nm) | Aniline blue (604nm) |
| **NINO Scientific: 1 (5.0%)** |  |  |  |  |  |  |
| Leishman<br>68 | No | No | Alcian blue (619nm) | Eosin Y/Eosin B<br>(518nm) | Neutral red (544nm) | Unknown dye (401nm) |
| **Ranjo : 1 (5.0%)** |  |  |  |  |  |  |
| Leishman<br>05 | No | No | Azure C (615nm) | Azocarmine G (512nm) | Neutral red (540nm) | Unknown dye (417nm) |
| **Searlie, UK: 1 (5.0%)** |  |  |  |  |  |  |
| Leishman<br>62 | **Yes** | **Yes** | Brilliant cresyl<br>blue(624nm) | Unknown dye (633nm) | Azure A(626nm) | Eosin Y/Eosin B (514nm) |
| **Trust Care Laboratories: 1 (5.0%)** |  |  |  |  |  |  |
| Leishman<br>156 | Yes | Yes | Sym-dimethyl thionine<br>(622nm) | Eosin B/Eosin Y<br>(516nm) | Phloxine B (549nm) | Unknown dye (375nm) |
| **Unknown Company: 1 (5.0%)** |  |  |  |  |  |  |
|  | No | No | Eosin Y/Eosin B (516nm) | Brilliant cresyl blue<br>(624nm) | Pyronine derivative<br>(560nm) | Azure A (626nm) |

**Table 4.**
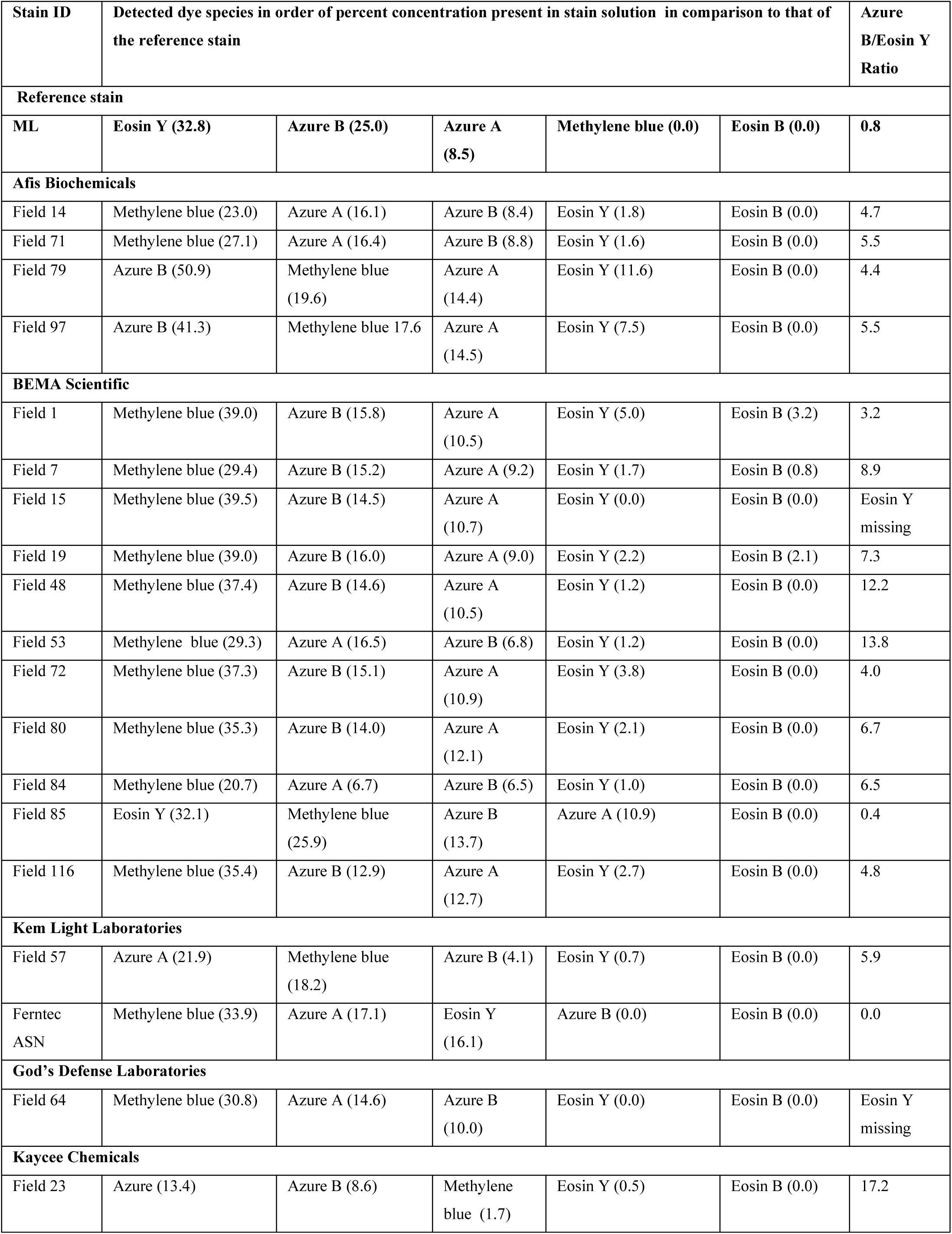
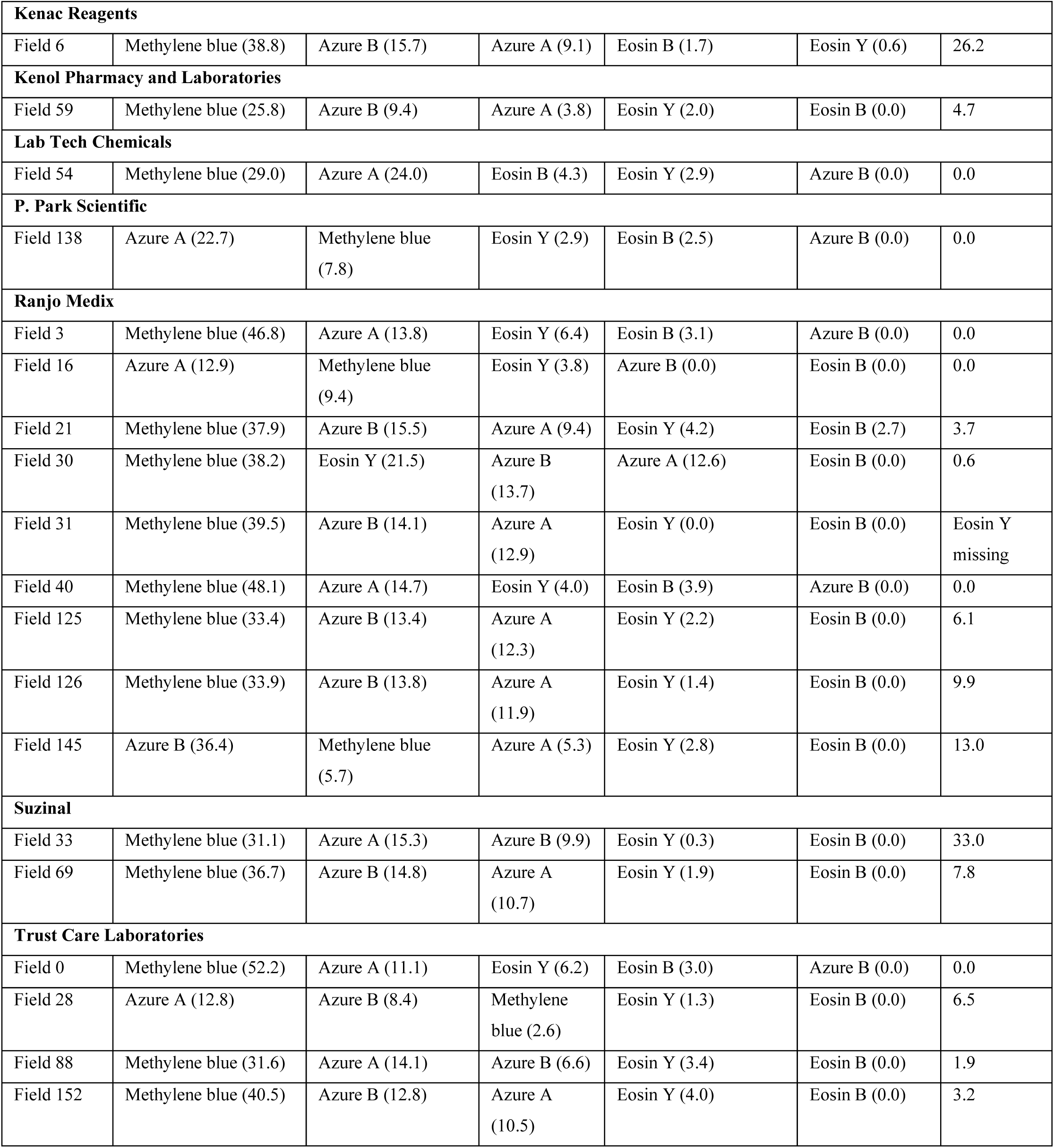
Identity and percent concentrations of dye species present in Field stains solution used for malaria microscopy in Plateau State Nigeria detected by high performance liquid chromatography (HPLC) assay.

**Table 5.** Identity and percent concentrations of dye species present in Giemsa stains solution used for malaria microscopy in Plateau State, Nigeria detected by high performance liquid chromatography (HPLC) assay.

| Stain ID | Detected dye species in order of percent concentration present in stain solution compared to that of the reference stain |  |  |  |  | Azure B:Eosin Y Ratio |
| --- | --- | --- | --- | --- | --- | --- |
| Reference stain |  |  |  |  |  |  |
| Marshall et al | Eosin Y (32.8) | Azure B (25.0) | Azure A (8.5) | Methylene blue (0.0) | Eosin B (0.0) | 0.8 |
| Afis Biochemicals |  |  |  |  |  |  |
| Giemsa 65 | Azure A (35.6) | Methylene blue (14.8) | Eosin B (10.9) | Eosin Y (9.5) | Azure B (4.4) | 0.5 |
| ANDi Laboratories, Lagos |  |  |  |  |  |  |
| Giemsa 00 | Azure B (21.0) | Eosin Y (18.8) | Azure A (7.0) | Methylene blue (0.0) | Eosin B (0.0) | 1.1 |
| Giemsa 132 | Azure A (20.5) | Azure B (15.8) | Eosin Y (8.4) | Eosin B (4.0) | Methylene blue (1.6) | 1.9 |
| Antec Diagnostics Ltd |  |  |  |  |  |  |
| Giemsa 120 | Azure B (28.1) | Azure A (13.0) | Eosin Y (10.8) | Methylene blue (2.2) | Eosin B (0.0) | 2.6 |
| Apex Laboratories Reagents |  |  |  |  |  |  |
| Giemsa 55 | Azure B (22.9) | Azure A (18.0) | Eosin Y (9.6) | Eosin B(4.4) | Methylene blue (4.0) | 2.4 |
| Giemsa 141 | Azure A(31.4) | Eosin Y (15.6) | Eosin B (5.3) | Azure B (3.5) | Methylene blue (0.0) | 0.2 |
| BEMA Scientific |  |  |  |  |  |  |
| Giemsa 4 | Eosin B (11.1) | Azure B (8.3) | Eosin Y (5.0) | Azure A (0.0) | Methylene blue (0.0) | 1.7 |
| Giemsa 20 | Azure B (26.1) | Eosin Y (21.5) | Eosin B (4.3) | Azure A (1.5) | Methylene blue (0.0) | 1.2 |
| Giemsa 22 | Azure A (33.5) | Eosin Y (15.9) | Azure B (13.4) | Methylene blue (0.0) | Eosin B (0.0) | 0.8 |
| Giemsa 48 | Azure A (38.9) | Eosin Y (15.9) | Azure B (9.7) | Eosin B (2.8) | Methylene blue (2.3) | 0.6 |
| Giemsa 97 | Azure A (20.5) | Azure B (9.4) | Eosin Y (6.0) | Methylene blue (2.5) | Eosin B (0.0) | 1.6 |
| Giemsa 107 | Azure A (42.2) | Eosin Y (11.5) | Azure B (9.5) | Eosin B (7.6) | Methylene blue (0.0) | 0.8 |
| Ferntec ASN |  |  |  |  |  |  |
| Giemsa 91 | Azure A (24.6) | Eosin Y (7.8) | Azure B (4.5) | Methylene blue (0.0) | Eosin B (0.0) | 0.6 |
| Hembarge Chemicals |  |  |  |  |  |  |
| Giemsa 154 | Methylene blue (32.1) | Eosin Y (11.9) | Eosin B (5.7) | Azure A (4.1) | Azure B (0.0) | 0.0 |
| Jos University Hospital Laboratories |  |  |  |  |  |  |
| Giemsa 2 | Azure B (36.5) | Eosin B (4.4) | Azure (3.6) | Eosin Y (1.5) | Methylene blue (1.5) | 24.3 |
| **Kaycee Chemicals** |  |  |  |  |  |  |
| Giemsa 84 | Azure A (40.4) | Eosin Y (11.8) | Azure B (5.6) | Methylene blue (1.7) | Eosin B (1.5) | 0.5 |
| **Kem Light Laboratories** |  |  |  |  |  |  |
| Giemsa 76 | Azure A (27.7) | Azure B (22.0) | Eosin Y (5.3) | Methylene blue (0.0) | Eosin B (0.0) | 4.2 |
| Giemsa 133 | Azure A (34.3) | Azure B (11.7) | Eosin Y (6.2) | Methylene blue (1.8) | Eosin B (0.0) | 1.9 |
| Giemsa 139 | Azure B (15.5) | Azure A (8.5) | Eosin Y (3.2) | Methylene blue (1.2) | Eosin B (0.8) | 4.8 |
| **Kermel** |  |  |  |  |  |  |
| Giemsa 121 | Methylene blue (29.4) | Azure A (27.5) | Eosin Y (15.0) | Azure B (0.0) | Azure B (0.0) | 0.0 |
| **Lab Tech Chemicals** |  |  |  |  |  |  |
| Giemsa 61 | Azure B (30.0) | Azure A (24.0) | Eosin Y (6.9) | Methylene blue (1.7) | Eosin B (0.0) | 4.3 |
| **Medicom** |  |  |  |  |  |  |
| Giemsa 8 | Azure A (23.7) | Eosin Y (23.6) | Eosin B (17.9) | Azure B (3.4) | Eosin B (0.0) | 0.8 |
| **Merck** |  |  |  |  |  |  |
| Giemsa 62 | Azure A (20.9) | Eosin B (17.1) | Eosin Y (6.8) | Azure B (2.2) | Methylene blue (0.0) | 0.3 |
| **Plateau State Specialist Hospital** |  |  |  |  |  |  |
| Giemsa 25 | Methylene blue (21.4) | Eosin Y (17.6) | Azure A (14.3) | Eosin B (13.0) | Azure B (0.0) | 0.0 |
| **Ranjo Medix** |  |  |  |  |  |  |
| Giemsa 117 | Azure A (29.4) | Methylene blue (16.6) | Azure B (15.7) | Eosin Y (7.4) | Eosin B (6.2) | 0.0 |
| Giemsa 142 | Azure A (25.8) | Eosin Y (15.1) | Eosin B (12.1) | Azure B (9.1) | Methylene blue (2.5) | 2.1 |
| Giemsa 142 | Methylene blue (31.6) | Azure A (14.1) | Azure B (6.6) | Eosin Y (3.4) | Eosin B (0.0) | 1.9 |
| **Raymond A. Lamb, London** |  |  |  |  |  |  |
| Giemsa 98 | Azure B (38.3) | Eosin Y (3.4) | Azure B (2.7) | Eosin B (0.5) | Methylene blue (0.5) | 0.8 |
| **S. Merck Chemicals** |  |  |  |  |  |  |
| Giemsa 199 | Azure B (22.9) | Eosin Y (21.3) | Azure A (6.0) | Methylene blue (3.1) | Eosin B (0.0) | 1.1 |
| **Trust Care Laboratories** |  |  |  |  |  |  |
| Giemsa 99 | Azure B (25.0) | Azure A (17.4) | Eosin Y (10.6) | Methylene blue (2.5) | Eosin B (0.0) | 2.4 |
| Giemsa 104 | Eosin B (20.8) | Azure A (20.6) | Eosin Y (10.8) | Azure B (10.4) | Methylene blue (0.0) | 1.0 |
| Giemsa 118 | Azure A (38.7) | Methylene blue (28.1) | Azure B (13.9) | Eosin B (3.1) | Eosin Y (1.3) | 10.7 |
| Giemsa 149 | Azure A (28.8) | Azure B (13.3) | Eosin Y (7.5) | Eosin B (3.4) | Methylene blue<br>(1.5) | 1.8 |
| Giemsa 150 | Azure A (22.7) | Azure B (20.0) | Eosin Y (13.9) | Eosin B (4.4) | Methylene blue<br>(1.1) | 1.4 |
| **Unknown manufacturer** |  |  |  |  |  |  |
| Giemsa 41 | Eosin Y (32.3) | Eosin B (30.3) | Azure A (17.4) | Methylene blue<br>(3.7) | Azure B (0.0) | 0.0 |
| Giemsa 157 | Azure A (24.9) | Azure B (22.7) | Eosin Y (10.1) | Eosin B (3.1) | Methylene blue<br>(2.0) | 2.2 |

**Table 6.**
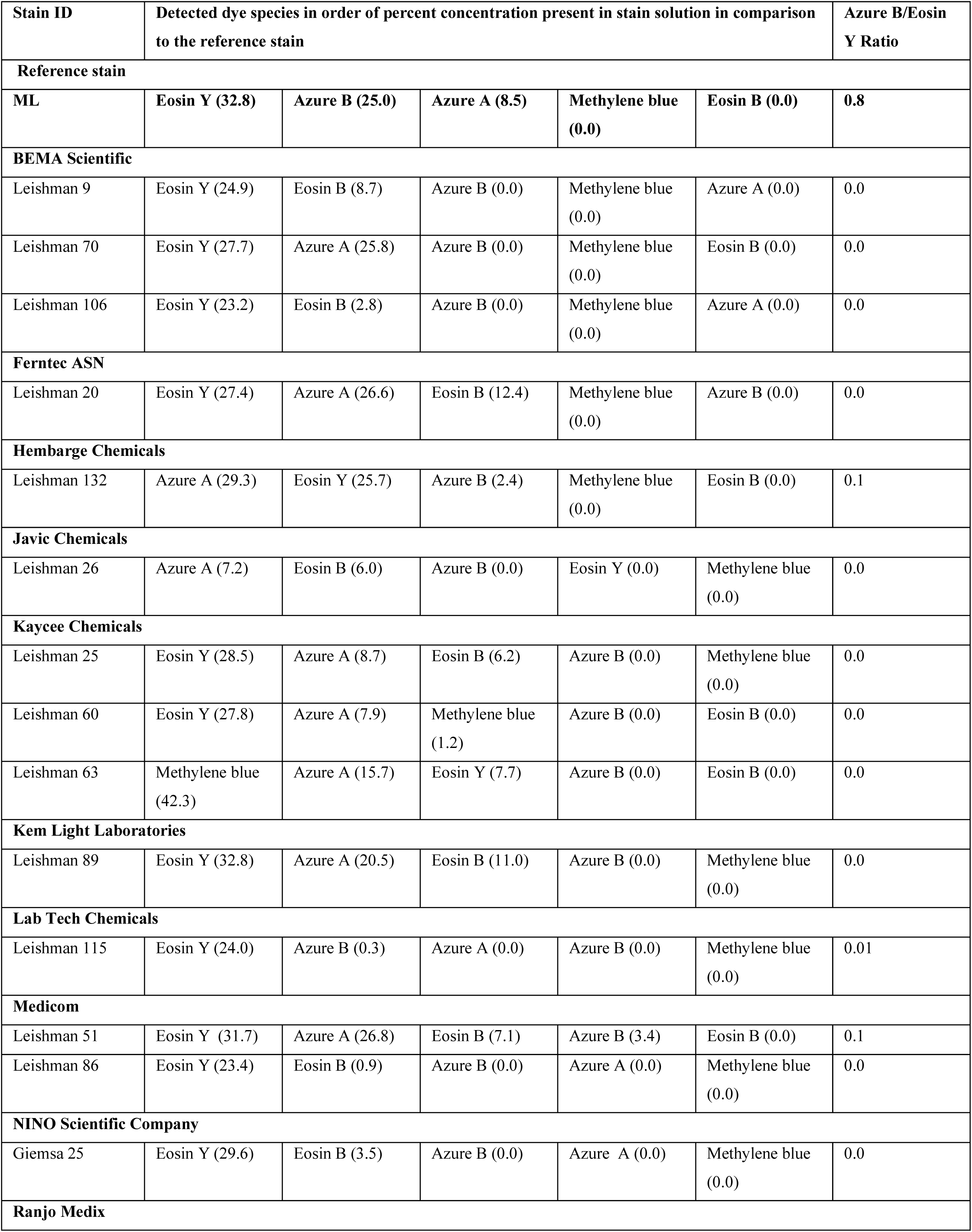
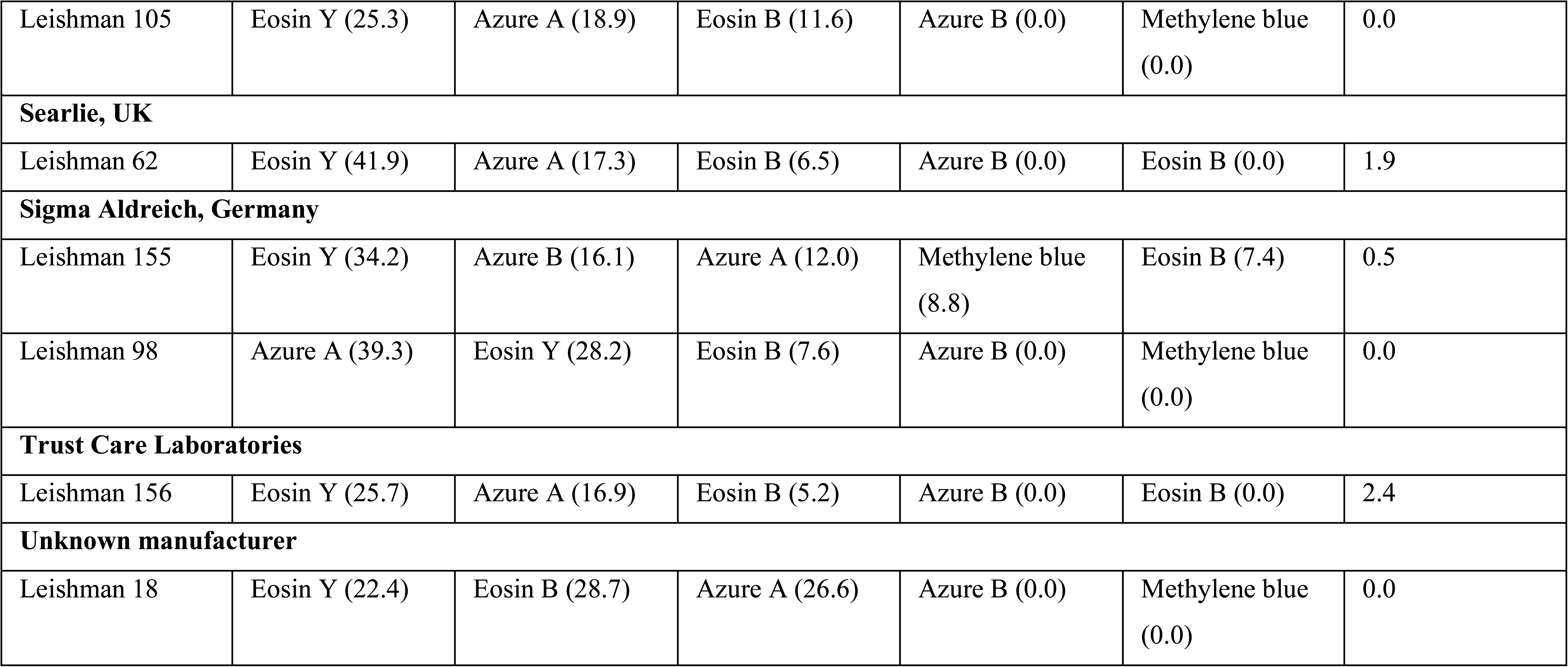
Identity and percent concentrations of dye species present in Leishman stains solution used for malaria microscopy in Plateau State Nigeria detected by high performance liquid chromatography (HPLC) assay.

