## Supplementary figures and images for "A multitude of dyes but unsatisfactory staining: Physico-chemical profile and factors associated with staining quality of Romanowsky-type stains used for malaria microscopy in Plateau State, Nigeria"

### Supplementary Fig 1.tiff

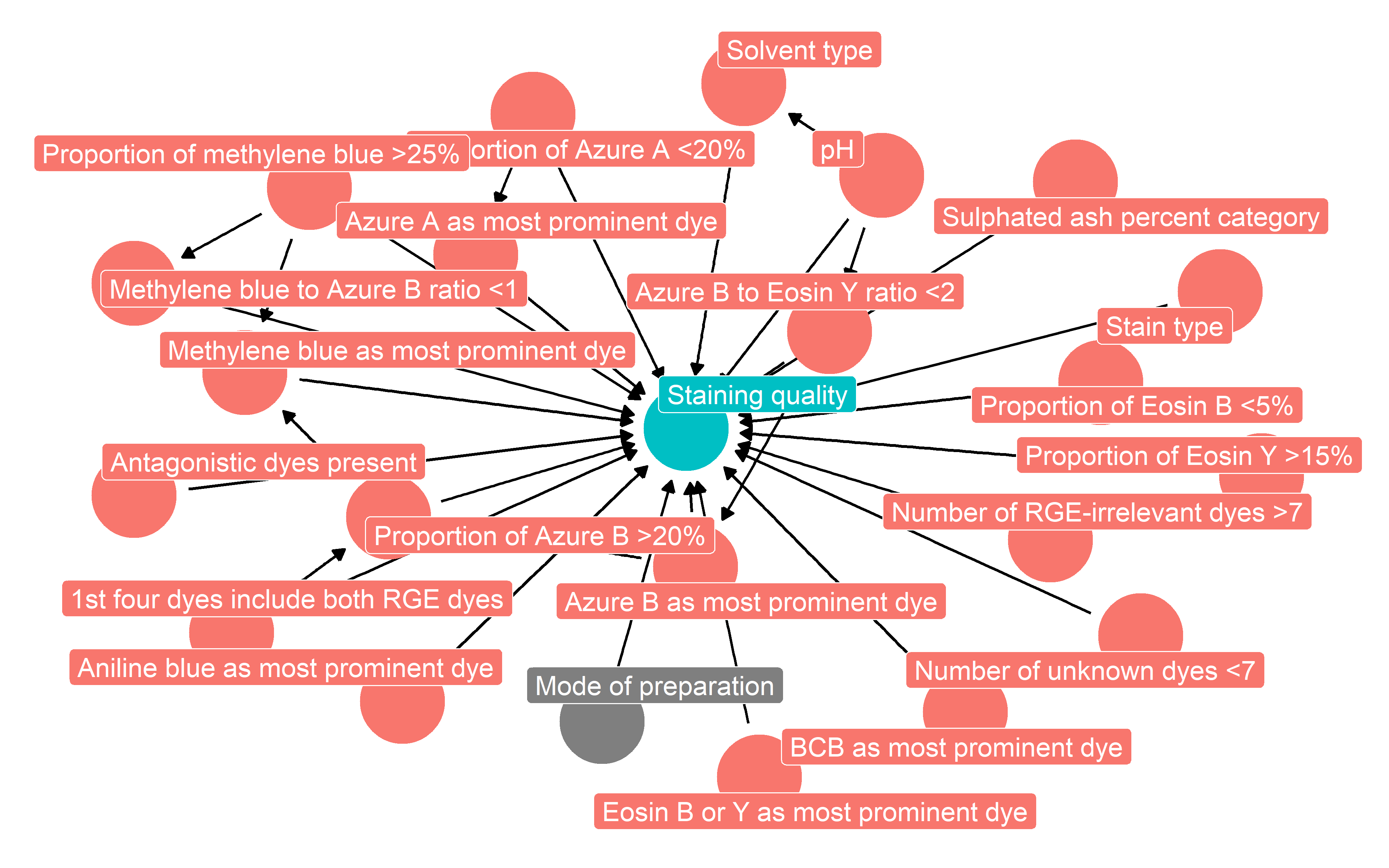

### Supplementary Fig 2.tiff

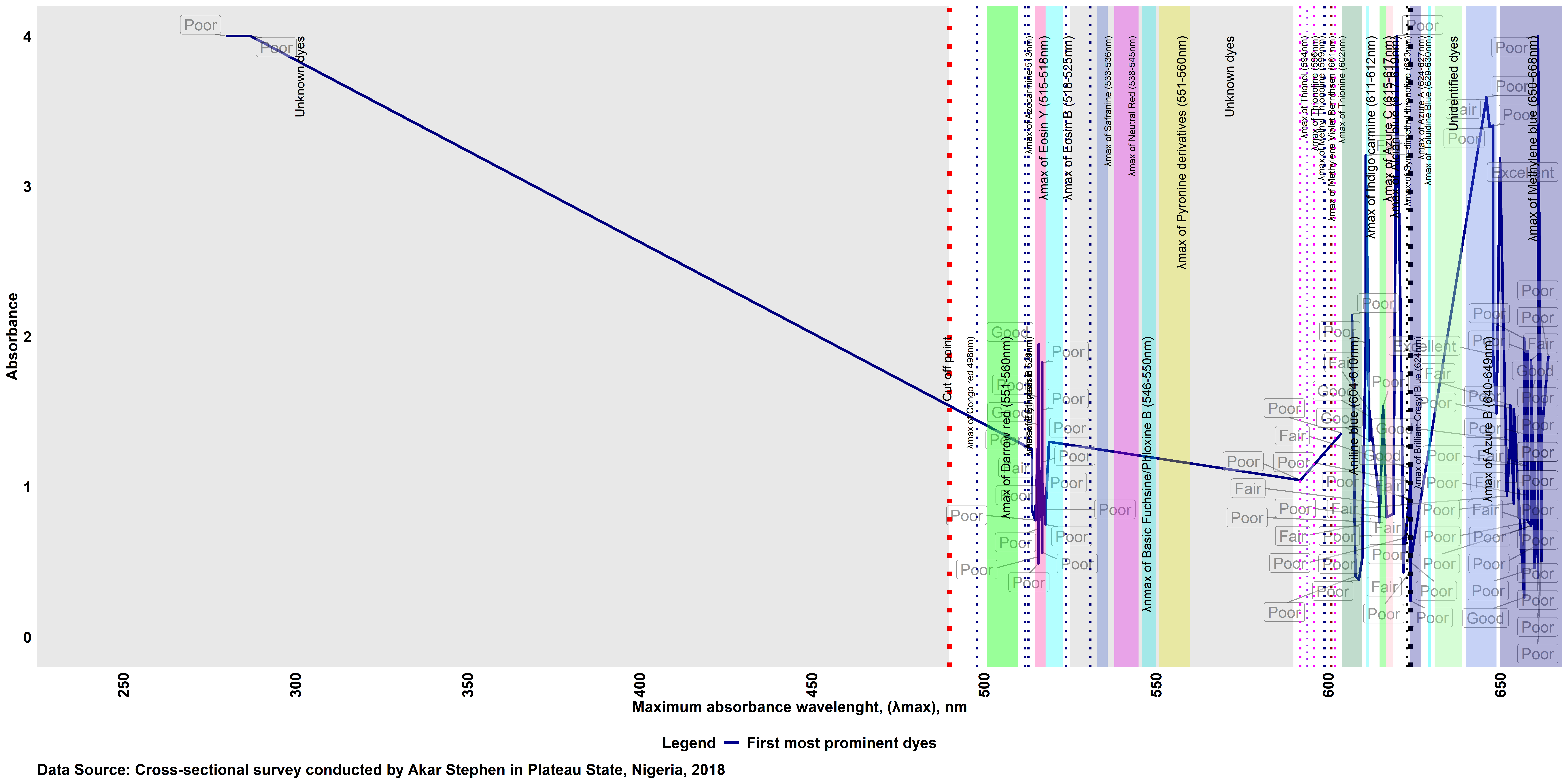

### Supplementary Fig 3.tiff

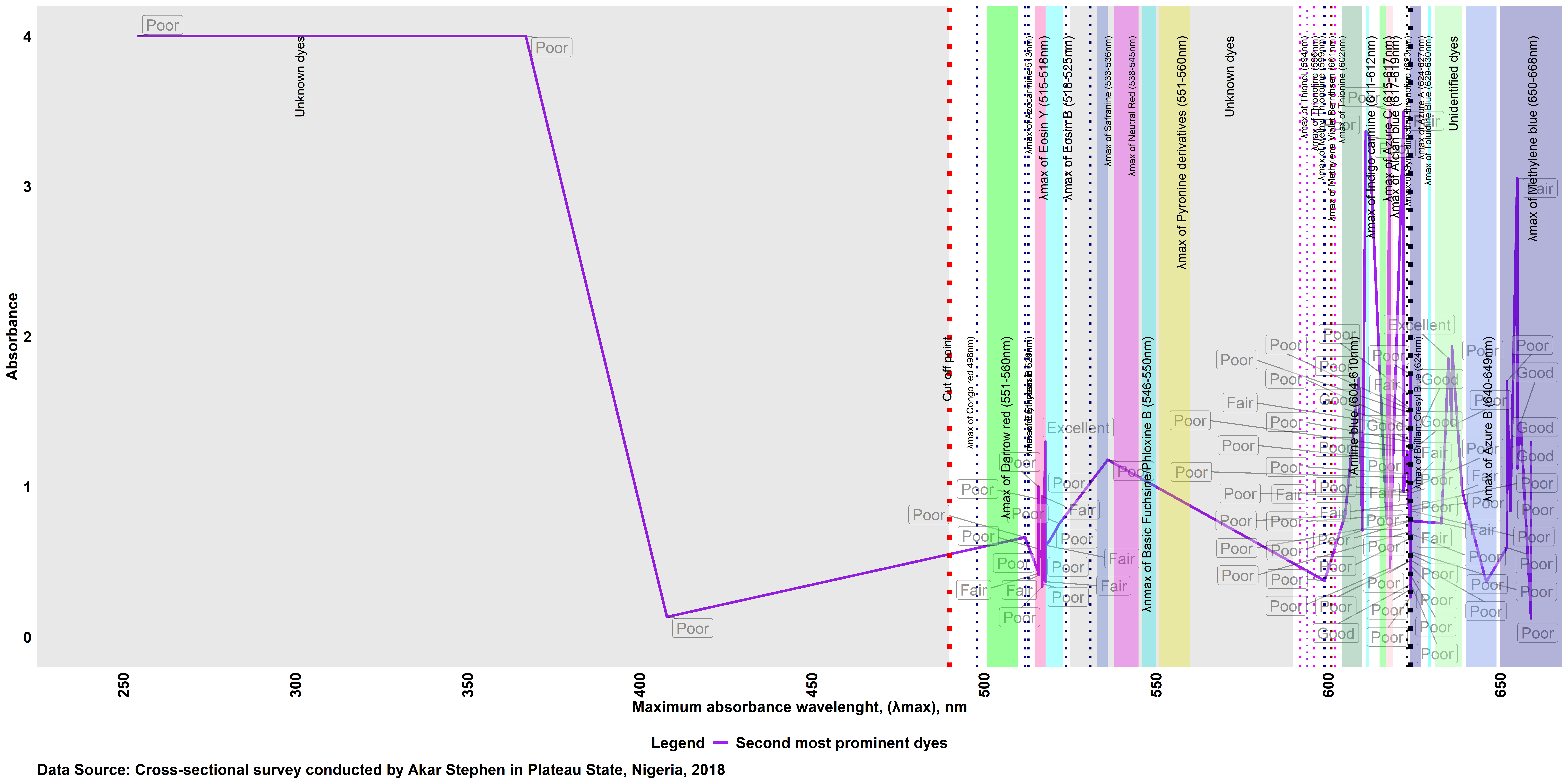

### Supplementary Fig 4.tiff

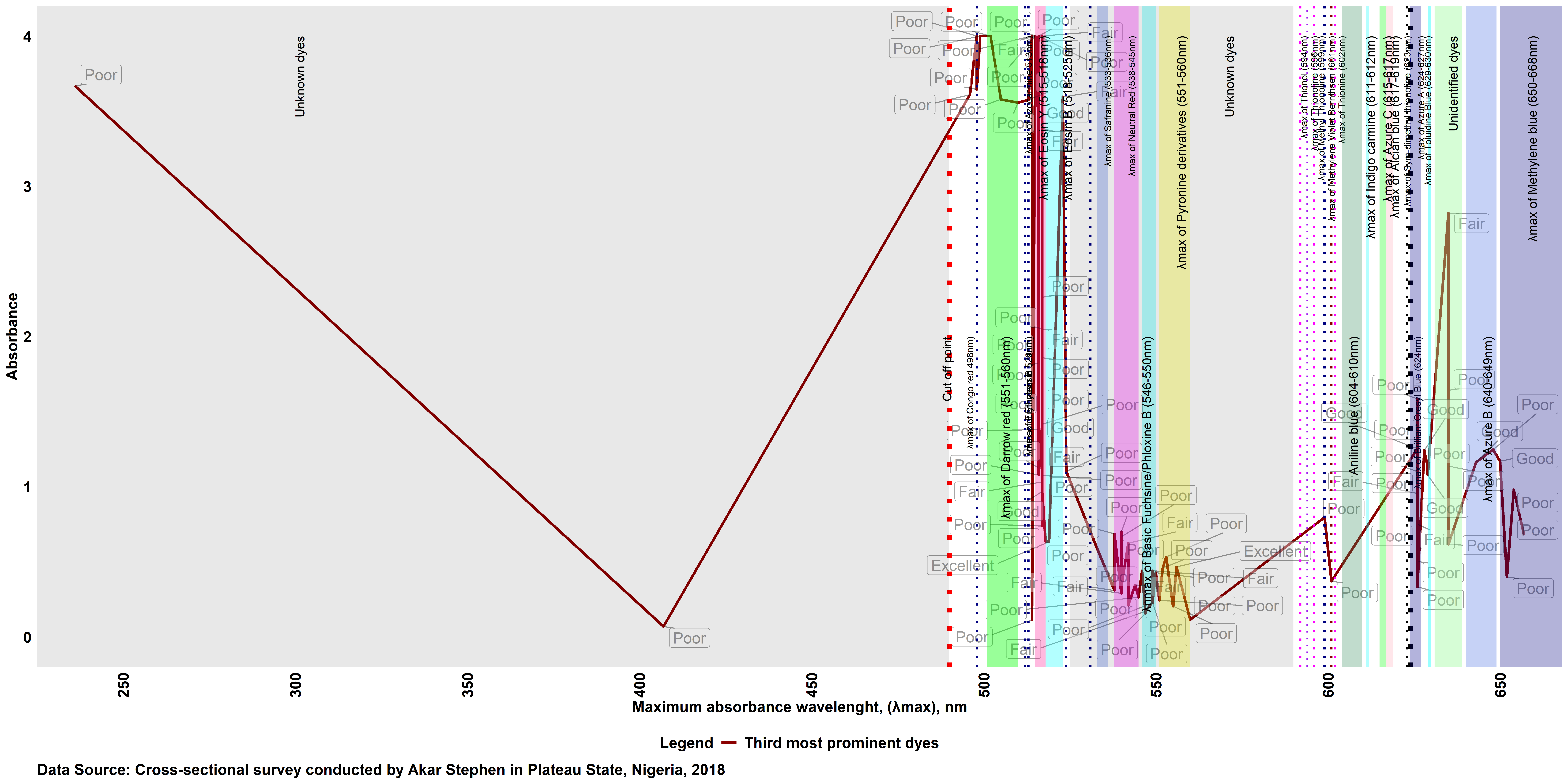

### Supplementary Fig 5.tiff

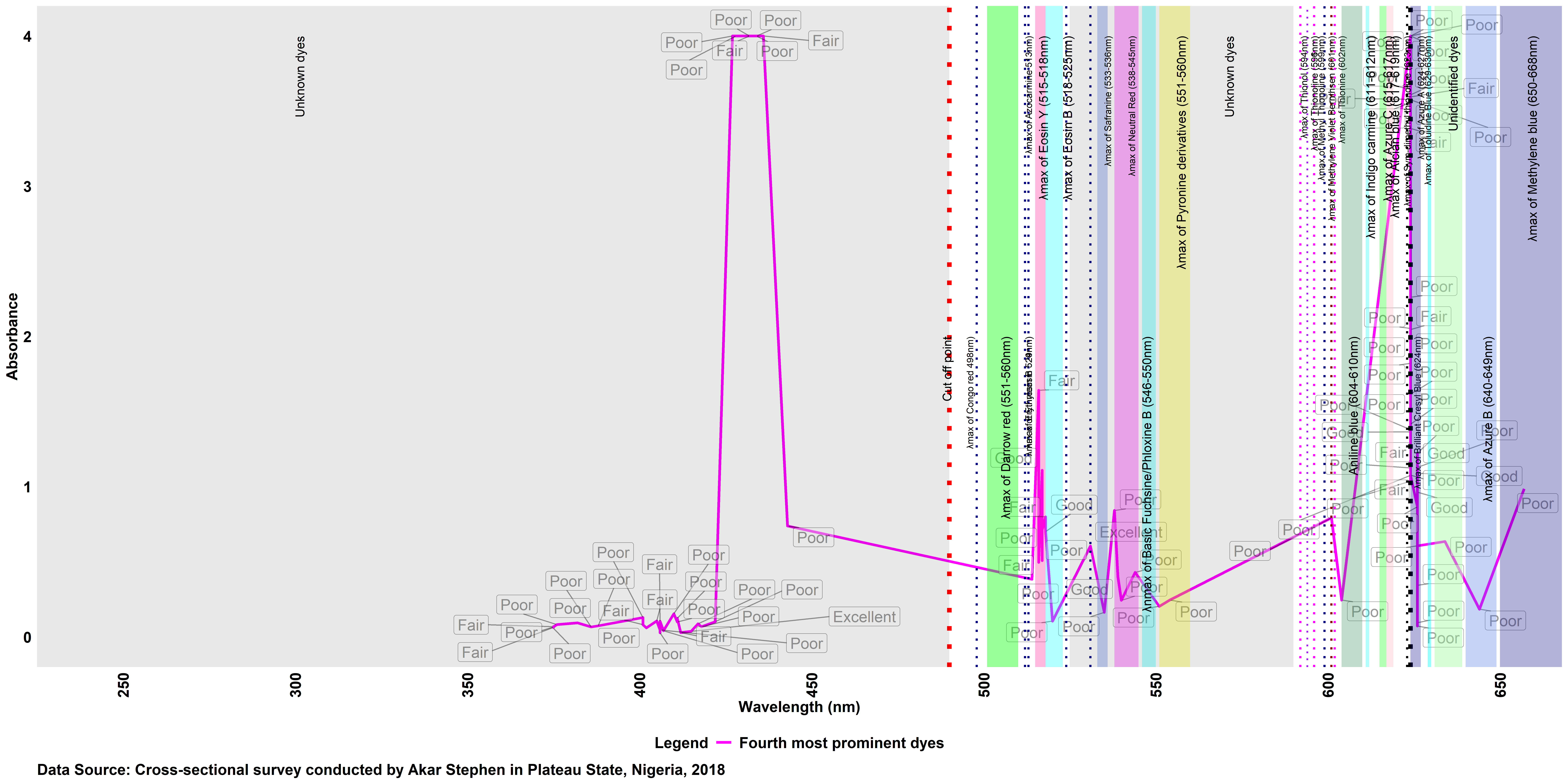
